# Progressive Supranuclear Palsy in India: Insights from a Large Multicenter Clinical Cohort (Project PAIR-PSP)

**DOI:** 10.64898/2026.01.25.26344786

**Authors:** Prashanth Lingappa Kukkle, Divyani Garg, Jacky Ganguly, Soaham Desai, Rukmini Mridula Kandadai, Sahil Mehta, Pettarusp Murzban Wadia, Deepika Joshi, Lulup Sahoo, Vijayashankar Paramanandam, Taallapalli Ashok Vardhan Reddy, Adreesh Mukherjee, Syam Krishnan, Kuldeep Shetty, Charulata Sankhla, Pankaj Agarwal, Heli S Shah, Suvorit Bowmick, Mitesh Chandarana, Malini Ayathu Venkat, Thenral S Geetha, Ramesh Menon, Sandeep Charugulla, Shakthivel Murugan, Ravi Gupta, Niraj Kumar, Atanu Biswas, Hrishikesh Kumar, Rupam Borgohain, Huw Morris, VL Ramprasad, Parkinson’s Research Alliance of India (PRAI) and the Global Parkinson’s Genetic Program (GP2)

## Abstract

**Background:** Progressive supranuclear palsy (PSP) is a rare and devastating tauopathy with limited global data. Given India’s large population, genetic diversity, and clinical heterogeneity, large multicenter datasets are crucial to enrich global understanding of PSP.

**Objective:** To characterize the demographic, clinical, and phenotypic profiles of a large multicenter Indian PSP cohort.

**Methods:** Subjects fulfilling MDS-PSP criteria were prospectively recruited across movement disorders centers (2021–2025). Standardized demographic and clinical data were collected.

**Results:** A total of 1,035 subjects were enrolled (M:F = 709:326), with a median age of 65 years and a mean onset age of 62.2±7.9 years. Regional distribution reflected pan-Indian recruitment (South 35%, North 26%, West 21%, East 18%). PSP-Richardson’s syndrome was most common (41%), followed by PSP-Parkinsonism (18%) and PSP-CBS (11%); rarer phenotypes included PSP-PI (7%), PSP-F (7%), PSP-PGF (5%), PSP-OM (2%), PSP-SL (1%), and PSP-C (1%). Falls occurred earliest in PSP-PGF (13.7 months) and PSP-SL (16.3 months), while PSP-P showed delayed disability (falls at 31 months) indicating progression patterns. Cognitive onset was prominent in PSP-F (21%) and PSP-SL (57%). Levodopa was prescribed to 893 patients; 186 (21%) reported >25% subjective benefit, and 358 (40%) reported ≤25% benefit. Amantadine was used in 351 (34%) patients, with improvement in 177.

**Conclusion:** This largest systematically profiled PSP cohort highlights both shared and distinctive features: high frequency of non-RS variants, aggressive course in PSP-RS/SL, better survival in PSP-P, and limited pharmacological benefit. These findings establish a foundation for longitudinal and genetic studies in diverse populations.

## Introduction

Progressive supranuclear palsy (PSP) is a primary tauopathy with diverse phenotypes and limited therapeutic options. Data from Indian tertiary centers highlight that atypical Parkinsonism contributes to nearly one-fifth of all movement disorder consultations, with PSP alone comprising about 10% of parkinsonian cases.(1) Over the past five decades, Indian researchers have steadily advanced the field through clinical descriptions, phenotypic characterizations, and genetic explorations. These works underscore both the heterogeneity of PSP manifestations and the urgent need for multicentric, population-based studies in India.(2–5) Globally, systematic reviews reveal striking geographic variation in PSP prevalence, subtypes, and prognosis, shaped by ancestry-related genetic factors, environmental exposures, healthcare access and limited data from underrepresented populations.(6)

However, most large-scale epidemiological and clinico-pathological studies originate from Europe and North America, leaving populations from the Global South grossly underrepresented.(6) Given India’s vast population and high frequency of the MAPT H1 haplotype, coupled with unique sociocultural and environmental contexts, large studies from this region are essential to refine diagnostic frameworks, delineate phenotype–genotype interactions, and enrich the global narrative of PSP and tauopathies.(6) This study represents a step toward that critical goal.

### Methodology

This study is part of an ongoing pan-India multicenter collaborative initiative conducted under the aegis of the Parkinson’s Research Alliance of India (PRAI), an amalgamation of eighteen movement disorders centers across the country. The project is being carried out in partnership with MedGenome Labs Pvt. Ltd. and is supported by the Global Parkinson’s Genetics Program (GP2), which is funded by the Aligning Science Across Parkinson’s Initiative, and which works with the implementation partner, the Michael J Fox Foundation for Parkinson’s Disease Research (MJFF).(7)

All participating centers obtained independent Institutional Ethics Committee/Review Board approvals for prospective patient recruitment. Individuals fulfilling the Movement Disorder Society (MDS) criteria for PSP were enrolled after providing written informed consent.(8) Diagnostic-certainty level and phenotypic designation were recorded separately: certainty was based on the highest level of MDS-PSP criteria fulfilled at recruitment, whereas phenotypic sub-classification reflected the predominant clinical syndrome. For phenotypic sub-classification, investigators classified cases according to the predominant presenting clinical syndrome; accordingly, rare atypical phenotypes not explicitly encompassed within the current MDS framework, such as PSP with predominant cerebellar dysfunction (PSP-C), were retained based on the basis of expert investigator assessment. Speech and language impairment was identified from routine clinical history and neurological examination at the participating centers, and phenotypic assignment was based on the MDS-PSP speech/language criterion (C1), including persistent nonfluent/agrammatic language disturbance and/or progressive apraxia of speech as assessed by the treating investigators. A uniform formal language battery was not applied across all centers. Recruitment involved systematic clinical evaluation, centralized data collection through a secure digital platform, and peripheral blood sampling for genetic analysis.(Appendix-1)

Data acquisition has been standardized across centers and includes detailed demographic profiles, clinical features, diagnostic categorization, and treatment-related information. Clinical profiling follows a harmonized protocol to ensure uniformity across sites. The biological samples are processed for genomic studies as part of the collaborative framework with MedGenome Labs and GP2. (Appendix-1)

All data are securely maintained under the PRAI consortium and are available for collaborative analyses upon request, in accordance with institutional and funding body guidelines.

## Results

### Demographics

From April 2021 to July 2025, 1,035 patients with PSP were enrolled (male: female = 709:326), with a median age of 65 years and a mean symptom onset at 62.2 ± 7.9 years (range: 38-87 years). The mean disease duration at recruitment was 39.3± 26.8 months (range : 2- 192 months). Participants were recruited from across India, with the largest contribution from the south (35%), followed by the north (26.1%), west (21.3%), and east (17.7%). (Figure-1A, 1B, Table-1)

**Figure-1:**
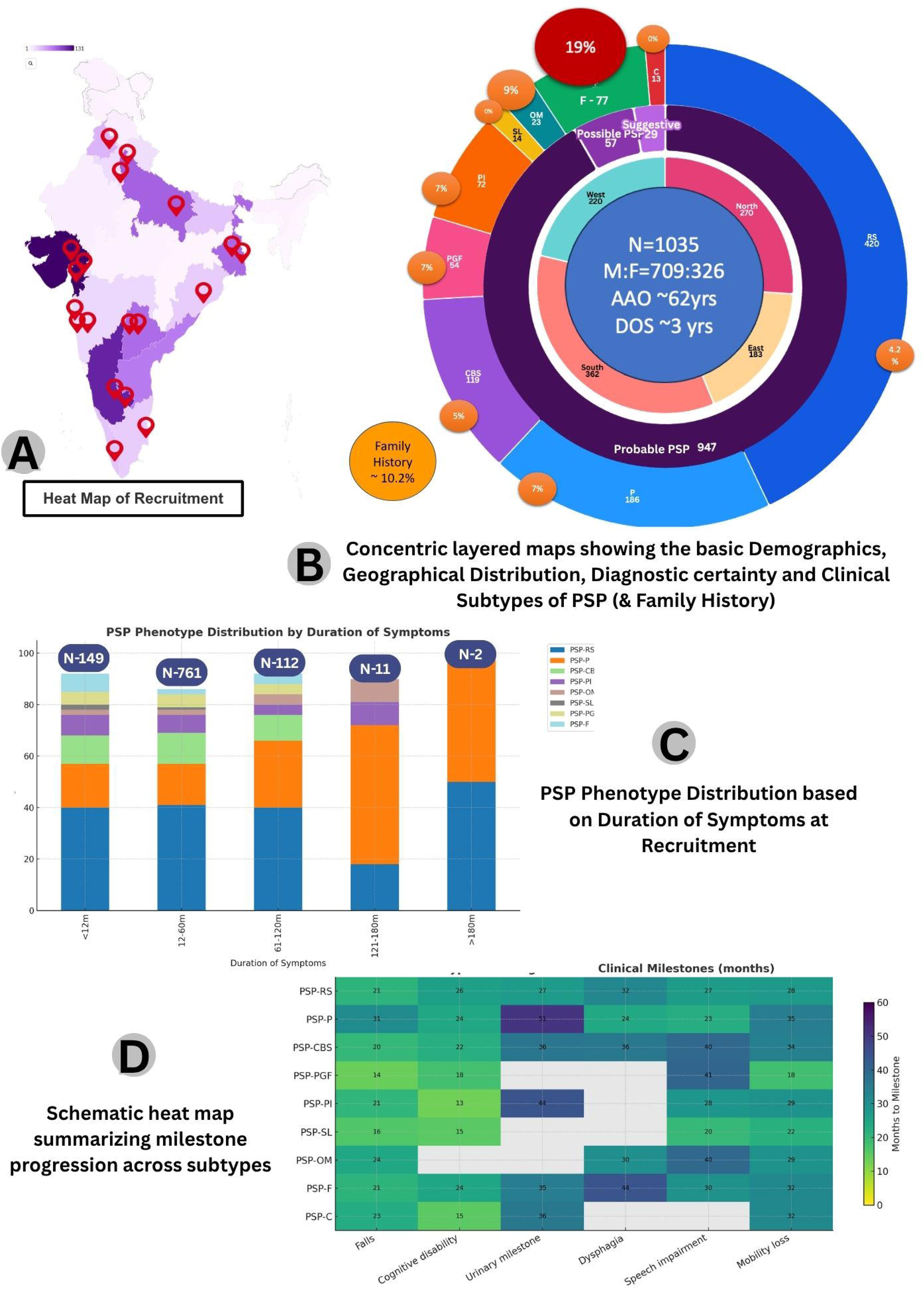
PAIR PSP Recruitment Infographic. 1A: Show the Heat map of recruitment of cases with locator marker showing the clinical centers . 1B: Layered concentric infographic with the center most circle showing the numbers of subjects (N), gender(M:F), age at onset (AAO) and duration of symptoms (DOS). The second concentric circle showing the the geographical distribution. The third concentric circle shows the number of subjects matching the MDS-PSP diagnostic criteria levels. The fourth (outermost) concentric circle shows the clinical phenotype distribution of the cases. The bubbles in the outermost concentric circle show the percentage of cases with family history of Parkinsonism. (RS : Richardson Syndrome, P : Parkinsonism, CBS - Corticobasal syndrome, PGF: Progressive gait freezing, PI: Postural Instability, SL: Speech Language, OM : Oculomotor, F: Frontal, C : Cerebellar) 1C: PSP Phenotype Distribution based up Duration of Symptoms at Recruitment 1D: Schematic heat map summarizing milestone progression across subtypes

**Table 1 :** Demographic profile of whole Cohort.

**Tables -1 Demographic profile of the whole cohort:**
|  | North Indian | East Indian | South Indian | West Indian | Total |
| --- | --- | --- | --- | --- | --- |
| No. of Subjects (%) | 270<br>(26.1) | 183<br>(17.7) | 362<br>(35) | 220<br>(21.3) | 1035 |
| Age at Onset<br>(Years, Range) | 59.5 ± 8.0 (38-87) | 62.5±7.9 (43-85) | 62.5±7.4 (41-80) | 64.6±8.1 (45-85) | 62.2±7.9 (38-87) |
| Male:Female<br>(Male-%) | 180:90 (66.7) | 132:51 (72.1) | 245:117 (67.7) | 142:78 (62.5) | 709:326 (68.5) |
| Duration of Symptoms<br>(months)<br>Mean ± SD (Range) | 36.6±25.4 (2-168) | 41.6±29.5 (2-192) | 40.7±26.1 (3-192) | 38.8±27.4 (4-144) | 39.3±26.8 (2-192) |

The cohort reflected India’s sociocultural diversity: most subjects were Hindu (87.1%) and represented all socioeconomic and educational strata. Consanguinity was reported in 6.7%, and a family history of Parkinsonism in 10.2%.

Diagnostic certainty, based on MDS-PSP criteria, classified the majority as Probable PSP (91.7%), with smaller proportions as Possible PSP (5.5%) and ‘Suggestive of PSP’ (2.8%). (Figure-1C)

Motor presentation was the most common initial clinical presentation (90%), followed by behavioral (5.4%) and cognitive (4.6%) presentations. The pattern of symptoms at onset varied, with the most frequent being falls (22.4%), slowness in daily activities (22.0%), and poor balance (17.1%) (Supplementary Figure A). Motor and non-motor symptoms documented during the disease course up to recruitment are shown in Supplementary Figures B and C, respectively.

Age at onset:

Age at onset exerted a clear influence on the demographic and clinical profile of PSP. (Table-2). The majority of cases clustered between 50–69 years, with a male predominance in all groups. Younger-onset patients had longer disease duration at recruitment and higher rates of parental consanguinity, whereas older-onset patients were more often from higher socioeconomic and educational strata. Two subjects had age at onset of 38 yrs, but have been on follow up for 2 and 5 years respectively and meeting Probable PSP criteria. These two have not been included in broader discussions. Motor symptoms were the predominant initial presentation across all ages, although behavioral and cognitive features were relatively more frequent in the elderly.

**Table 2 :** Age of Onset Based Clinical Profile of PSP subjects.

| Age at Onset | 40-49<br>years | 50-59<br>years | 60-69<br>years | 70-79<br>years | 80-89<br>years |
| --- | --- | --- | --- | --- | --- |
| **Demographic Features** |  |  |  |  |  |
| No. of Subjects | 53 | 331 | 451 | 184 | 14 |
| Male:Female | 31:22 | 206:125 | 317:134 | 143:41 | 11:3 |
| Mean age at onset(years)<br>(Range) | 46.1±2.6<br>(40-49) | 55.5±2.7<br>(50-59) | 64.1±2.8<br>(60-69) | 72.9±2.7<br>(70-79) | 81.6±2.3<br>(80-87) |
| Duration of symptoms<br>(months)<br>Mean ± SD(Range) | 51.3±36.4<br>(4-180) | 39.7±26.0<br>(2-144) | 40.5±28.6<br>(2-192) | 33.5±18.8<br>(3-96) | 26.6±16.6<br>(3-60) |
| Age at study inclusion (years)<br>Mean ± SD (Range) | 50.6±4.6<br>(42-67) | 58.9±3.6<br>(50-73) | 67.5±3.8<br>(56-83) | 75.6±3.0<br>(70-84) | 82.5±3.9<br>(71-88) |
| Education (≥graduate) | 10 (19%) | 96 (29) | 156 (35) | 81 (44) | 4 (29) |
| Economic Status (Upper class)<br>(%) | 6 (11) | 61(18) | 123 (27) | 61 (33) | 5 (36) |
| Consanguineous parentage (%) | 4 (8) | 23 (7) | 27 (6) | 15 (8) | 0 (0) |
| Family history of Parkinsonism<br>(%) | 4 (8) | 24 (7) | 52 (11) | 23(12) | 2 (14) |
| **Initial Symptom Category** |  |  |  |  |  |
| Behavioral (%) | 2 (4) | 3 (1) | 21 (5) | 9 (5) | 1 (7) |
| Cognitive (%) | 2 (4) | 11 (3) | 16 (4) | 13 (7) | 0 (0) |
| Motor (%) | 49 (92) | 294 (89) | 411(91) | 161 (88) | 13 (93) |
| **Diagnostic Certainty of PSP (MDS-PSP)** |  |  |  |  |  |
| Probably PSP (%) | 50 (94) | 313 (95) | 412 (91) | 158 (86) | 14 (100) |
| Possible PSP (%) | 1 (2) | 11(3) | 27 (6) | 18 (10) | 0 (0) |
| Suggestive of PSP (%) | 2 (4) | 7 (2) | 12 (3) | 8 (4) | 0 (0) |

| Primary Clinical Phenotype |  |  |  |  |  |
| --- | --- | --- | --- | --- | --- |
| PSP-RS (%) | 22 (41) | (45) | 177 (39) | 66 (36) | 6 (43) |
| PSP-P (%) | 9 (17) | 53 (16) | 80 (18) | 41(22) | 3 (21) |
| PSP-CBS (%) | 4 (8) | 35 (11) | 53 (12) | 25 (14) | 1 (7) |
| PSP-PI (%) | 5 (9) | 20 (6) | 35 (8) | 12 (6) | 0 (0) |
| PSP -OM (%) | 4 (8) | 8 (2) | 9 (2) | 1 (1) | 0 (0) |
| PSP-SL (%) | 0 (0) | 2 (1) | 7 (2) | 5 (3) | 0 (0) |
| PSP-PGF (%) | 1 (2) | 18 (5) | 23 (5) | 11(6) | 1 (7) |
| PSP-F (%) | 5 (9) | 22 (7) | 35 (8) | 11 (6) | 2 (14) |

- ❑ Two subjects had age of onset of 38 years and are not reflected in this table. They had been on follow up for 2 yrs and 5 yrs respectively meeting the 'probably PSP' criteria at recruitment with prominent vertical gaze palsy and postural instability. - ❑ Percentages are shown in the parenthesis and indicate in relation to specific subgroups (rounded to nearest whole number)

While “Probable PSP” was the dominant diagnostic category, its certainty decreased with later onset, accompanied by a rise in “Possible PSP.” Phenotypically, PSP-Richardson’s syndrome was most common across groups, but its relative frequency declined with advancing age at onset, while PSP-Parkinsonism and PSP-Corticobasal phenotypes increased among older cohorts. (Table-2)

### MDS-PSP Certainty criteria

Analysis of MDS-PSP diagnostic features across age groups revealed distinct trends. Vertical gaze palsy was the most frequent oculomotor finding, but its prevalence declined with older onset (75% to 57% change), whereas slow saccades became more prominent with advancing age. Similarly, early unprovoked falls were more typical of younger onset PSP (40-49 yrs group: P1-77%, P2- 7%), while pull-test abnormalities were increasingly recognized in older cohorts (70-79 yrs group: P1- 61%, P2: 21%).

Across all groups, akinetic-rigid, levodopa-resistant Parkinsonism remained the dominant motor feature. Cognitive and speech/language abnormalities were present in about one-third of patients, with a tendency toward higher frequency in younger and very elderly onset, suggesting a probable bimodal distribution. Although “Probable PSP” accounted for the majority in all groups, diagnostic certainty decreased with later onset. Phenotypically, PSP-Richardson’s was the most common, but its proportion declined with age, while PSP-Parkinsonism and PSP-Corticobasal variants increased in older subjects. (Table-3, Figure-1B)

**Table 3 :** Age groups based MDS-PSP diagnostic certainty criteria.

| **Age at Onset** | **40-49**<br>**years** | **50-59**<br>**years** | **60-69**<br>**years** | **70-79**<br>**years** | **80-89**<br>**years** |
| --- | --- | --- | --- | --- | --- |
| No. of Subjects | 53 | 331 | 451 | 184 | 14 |
| **<u>Mandatory Criteria (B1)</u>** |  |  |  |  |  |
| Sporadic occurrence | 49 | 320 | 431 | 175 | 13 |
| **<u>Core Clinical Features - Oculomotor</u>** |  |  |  |  |  |
| O1 – Vertical Supranuclear gaze Palsy (%) | 40 (75) | 238 (72) | 307 (68) | 103 (56) | 8 (57) |
| O2 – Slow Velocity of Vertical Saccades (%) | 10 (19) | 77 (23) | 116 (26) | 60 (33) | 5 (36) |
| O3 – Frequent Macro Square wave jerks or<br>Eyelid opening apraxia (%) | 2 (4) | 10 (3) | 20 (4) | 12 (6) | 0 (0) |
| **<u>Core Clinical Features – Postural Instability</u>** |  |  |  |  |  |
| P1 – Repeated Unprovoked falls within 3<br>years (%) | 41 (77) | 242 (73) | 308 (68) | 112 (61) | 8 (57) |
| P2 - Tendency to fall on pull test within 3<br>years(%) | 4 (7) | 57 (17) | 72 (16) | 38 (21) | 5 (36) |
| P3 – More than two steps backwards on the<br>pull test (%) | 0 (0) | 13 (4) | 36 (8) | 16 (9) | 1 (7) |
| **<u>Core Clinical Features – Akinesia</u>** |  |  |  |  |  |
| A1 – Progressive gait freezing within 3 years<br>(%) | 11 (21) | 99 (30) | 115 (25) | 50 (27) | 3 (21) |
| A2 – Parkinsonism, akinetic-rigid,<br>predominantly axial and levodopa<br>resistant(%) | 29 (55) | 169 (51) | 213 (47) | 83 (45) | 9 (64) |
| A3 – Parkinsonism, with tremor and /or<br>asymmetric and /or levodopa responsive<br>(%) | 7 (13) | 29 (9) | 71 (16) | 25 (14) | 2 (14) |

| **Core Clinical Features – Cognitive Dysfunction** |  |  |  |  |  |
| --- | --- | --- | --- | --- | --- |
| C1 – Speech / Language disorder (%) | 20 (38) | 117 (35) | 139 (31) | 61 (33) | 4 (29) |
| C2 – Frontal cognitive / behavioral presentation (%) | 20 (38) | 98 (30) | 128 (28) | 47 (25) | 7 (50) |
| C3 – Corticobasal syndrome (%) | 6 (11) | 39 (12) | 61 (13) | 23 (12) | 0 (0) |
| **Diagnostic Certainty of PSP (MDS-PSP)** |  |  |  |  |  |
| Probably PSP (%) | 50 (94) | 313 (95) | 412 (91) | 158 (86) | 14(100) |
| Possible PSP (%) | 1 (2) | 11 (3) | 27 (6) | 18 (10) | 0(0) |
| Suggestive of PSP (%) | 2 (4) | 7 (2) | 12 (3) | 8 (4) | 0 (0) |
| **Primary Clinical Phenotype** |  |  |  |  |  |
| PSP-RS (%) | 22 (41) | 149 (45) | 177 (39) | 66 (36) | 6 (43) |
| PSP-P (%) | 9 (17) | 53 (16) | 80 (18) | 41(22) | 3 (21) |
| PSP-CBS (%) | 4 (7) | 35 (11) | 53 (12) | 25 (14) | 1 (7) |
| PSP-PI (%) | 5 (9) | 20 (6) | 35 (8) | 12 (6) | 0 (0) |
| PSP -OM (%) | 4 (7) | 8 (2) | 9 (2) | 1 (1) | 0 (0) |
| PSP-SL (%) | 0 (0) | 2 (1) | 7 (2) | 5 (3) | 0(0) |
| PSP-PGF (%) | 1 (2) | 18 (5.4) | 23 (5) | 11(6) | 1 (7) |
| PSP-F (%) | 5 (9) | 22 (7) | 35 (8) | 11 (6) | 2 (14) |

- ☐ Two subjects had age of onset of 38 years and are not reflected in this table. They had been on follow up for 2 yrs and 5 yrs respectively meeting the ‘probably PSP’ criteria at recruitment with prominent vertical gaze palsy and postural instability. - ☐ Percentages indicate in relation to specific subgroups (rounded to nearest whole number)

### Duration of Disease

When stratified by disease duration, most patients (88%) fell within the 12–60 month group, while very long survivors beyond 10 years were rare (1%). (Table-4, Figure-1C) Younger age at onset and higher socioeconomic/educational status were more common among patients with longer disease duration. Motor onset dominated across all groups, with behavioral and cognitive presentations remaining infrequent. Diagnostic certainty was highest in intermediate disease durations, while very early and very long disease durations showed greater proportions of “Possible” or “Suggestive” PSP. Phenotypically, PSP-Richardson’s syndrome remained the most common subtype overall but declined sharply in longer-duration groups, whereas PSP-Parkinsonism increased progressively with duration, comprising over half of cases in the >10-year survivors. Corticobasal presentations peaked at intermediate durations and were absent among long survivors. These trends suggest that phenotype and baseline demographics influence disease course and survival in PSP. (Table-4)

**Table 4 :** Clinical Profile of PSP by Duration of Symptoms:

| **Duration of Symptoms** | **<12**<br>**Months** | **12-60**<br>**Months** | **61-120**<br>**Months** | **121-180**<br>**Months** | **>180**<br>**Months** |
| --- | --- | --- | --- | --- | --- |
| **<u>Demographic Features</u>** |  |  |  |  |  |
| No. of Subjects | 149 | 761 | 112 | 11 | 2 |
| Male:Female | 104:45 | 519:242 | 77:35 | 8:3 | 1:1 |
| Mean age at onset, years<br>(Range) | 62.6±8.3<br>(41-87) | 62.4±7.9<br>(38-85) | 60.1±8.0<br>(41-78) | 58.1±6.6<br>(45-63) | 62.5±0.7<br>(62-63) |
| Duration of symptoms, months<br>(Range) | 10.1±2.9<br>(2-12) | 36.1±13.4<br>(13-60) | 86.1±17.6<br>(62-120) | 153.8±15.9<br>(132-180) | 192 |
| Age at study inclusion years<br>(Range) | 63.5±8.2<br>(43-83) | 65.5±7.8<br>(40-88) | 67.1±7.9<br>(50-84) | 70.9±6.1<br>(58-76) | 78<br>(78-78) |
| Education (≥graduate) | 42 (28) | 258 (34) | 43 (38) | 5 (45) | 0 (0) |
| Economic Status (Upper class) | 24 (16) | 183 (24) | 42 (37) | 6 (54) | 1 (50) |
| **<u>Initial Symptom Category</u>** |  |  |  |  |  |
| Behavioral (%) | 5 (3) | 46 (6) | 5 (4) | 0 (0) | 0 (0) |
| Cognitive (%) | 3 (2) | 35 (5) | 4 (4) | 0 (0) | 0 (0) |
| Motor (%) | 141 (95) | 675 (89) | 101(90) | 11 (100) | 2 (100) |
| **<u>Diagnostic Certainty of PSP (MDS-PSP)</u>** |  |  |  |  |  |
| Probably PSP (%) | 130 (87) | 704 (92) | 104 (93) | 9 (82) | 2 (100) |
| Possible PSP (%) | 12 (8) | 41 (5) | 3 (3) | 1 (9) | 0 (0) |
| Suggestive of PSP (%) | 7 (5) | 16 (2) | 5 (4) | 1 (9) | 0 (0) |
| **<u>Primary Clinical Phenotype</u>** |  |  |  |  |  |
| PSP-RS (%) | 60 (40) | 312 (41) | 45 (40) | 2 (18) | 1 (50) |
| PSP-P (%) | 26 (17) | 124 (16) | 29 (26) | 6 (54) | 1 (50) |
| PSP-CBS (%) | 17 (11) | 91 (12) | 11 (10) | 0 (0) | 0 (0) |
| PSP-PI (%) | 12 (8) | 55 (7) | 4 (4) | 1(9) | 0 (0) |
| PSP -OM (%) | 3 (2) | 14 (2) | 4 (4) | 1 (9) | 0 (0) |
| PSP-SL (%) | 3 (2) | 11 (1) | 0 (0) | 0 (0) | 0 (0) |
| PSP-PGF (%) | 8 (5) | 42 (5) | 4 (4) | 0 (0) | 0 (0) |
| PSP-F (%) | 10 (7) | 16 (2) | 5 (4) | 0 (0) | 0 (0) |

- Two subjects had age of onset of 38 years and are not reflected in this table - Percentages are shown in parenthesis and indicate in relation to specific subgroups

### Subtype-wise Clinical Profiles of PSP

***1. PSP-Richardson’s Syndrome (PSP-RS):*** PSP-RS was the most frequent phenotype in the cohort (n=420; 40% of all cases). (Table-5) Subjects had a slightly younger mean age at onset (61.6 years) as compared to whole group, with the majority presenting with motor onset symptoms (92%). Diagnostic certainty was highest among all subtypes, with 97% fulfilling “Probable PSP” criteria, underscoring its classical presentation and relatively straightforward recognition. Falls appeared early (mean 21 months), and severe mobility dependence followed within 28 months, reflecting its more aggressive natural history. Regarding severity, often subjects were graded ‘moderately ill’ (26%) or ’markedly to severely ill’ (24%) by clinical global impression (CGI) scores. Consanguinity (9.5%) and family history (4%) were relatively uncommon but present. Regionally, PSP-RS was broadly distributed, with higher representation from South India (40%). Compared with other subtypes, PSP-RS declined in frequency in long-survivor groups (Table-4), confirming its poorer survival profile compared to other subtypes.
***2. PSP-Parkinsonism (PSP-P):*** PSP-P (n=186; 18%) represented the second largest group and differed from PSP-RS in several respects. (Table-5) The mean age at onset (62.9 years) was slightly later, and the mean duration of symptoms (46 months) was longer, consistent with slower disease progression. Motor onset was nearly universal (95%), but the proportion of “Probable PSP” (90%) was lower than PSP-RS, with higher fractions categorized as “Possible” or “Suggestive” (10%). Family history was more frequent (7%). Regional distribution was balanced across India. Falls and cognitive decline occurred later than in PSP-RS (31 and 24 months, respectively), urinary dependence was delayed (mean 50 months), as was mobility loss (35 months). The slower tempo was reflected in CGI severity, which predominated in the ‘moderate/mild’ range. Importantly, as seen in Table-4, PSP-P proportionately increased in long duration cohorts (comprising more than half of the >10-year survivors), highlighting its relative survival advantage.
***3. PSP-Corticobasal Syndrome (PSP-CBS):*** PSP-CBS (n=119; 12%) was motor-dominant (94%) with lower diagnostic certainty (88% “Probable PSP”). (Table-5) Falls and cognitive decline occurred around 20–22 months, close to PSP-RS, but dysphagia and speech decline showed a later onset (∼36–40 months), suggesting asymmetric disability accrual. Urinary dependence was also relatively early (∼36 months). While CBS patients survived longer than PSP-RS, this was rarely beyond 10 years, marking it as an intermediate-aggressiveness phenotype.
***4. PSP-Progressive Gait Freezing (PSP-PGF):*** PSP-PGF (n=54; 5%) was defined by 100% motor onset with early and rapidly progressive gait freezing. (Table-5) Mean age at onset was 63 years, with intermediate duration (36 months). Falls occurred earliest of all subtypes (∼14 months), and mobility dependence followed at ∼18 months, underscoring its early axial disability. However, cognitive decline and urinary complications were relatively infrequent. Diagnostic certainty was lower (83% “Probable PSP”), reflecting overlap with other parkinsonian syndromes. Survival did not extend into long-duration groups, suggesting a shorter but gait-dominant course.
***5. PSP-Postural Instability (PSP-PI):*** PSP-PI (n=72; 7%) shared features with PSP-PGF, showing frequent early falls and imbalance.(Table-5, Supplementary Table S1) Age at onset was 62 years, and the mean disease duration was 36 months. Falls milestone occurred at ∼21 months, cognitive disability even earlier (∼13 months), and mobility loss around 29 months. This combination of early cognitive and axial impairment distinguished PSP-PI from PSP-P. CGI ratings were skewed toward “markedly ill,” and regional clustering was noted in South India. Its progression was faster than PSP-P but slower than PSP-RS, placing it in an intermediate survival category.
***6. PSP-Speech/Language (PSP-SL):*** Though rare (n=14; 1.4%), PSP-SL was highly distinctive. (Table-5) Patients had the oldest onset (67 years), shortest mean disease duration (26 months), and extreme male predominance (92%). Non-motor onset was striking: 57% cognitive and language symptoms. Falls occurred early (∼16 months), speech impairment evolved fastest (∼20 months), and cognitive disability followed within 15 months. These patients progressed rapidly to severe illness, with few surviving beyond 4 years. PSP-SL thus represents one of the most aggressive non-motor variants of PSP. The PSP-SL subgroup warrants cautious interpretation because of the small number of cases, and the observed regional distribution broadly mirrored the overall cohort recruitment pattern rather than establishing a clear geographic signal.
***7. PSP-Ocular Motor (PSP-OM):*** PSP-OM (n=23; 2%) presented at an earlier age (mean 58 years) and was unique for its near equal gender distribution (M:F = 16:17), contrasting with the male predominance seen in most subtypes. Oculomotor features were prominent at onset (91% motor-dominant but with characteristic eye movement abnormalities). Diagnostic certainty was moderate (87% “Probable PSP”). Regionally, it showed a mixed spread. This subtype appeared across different age at onsets, without better survival, suggesting a distinct but not long-lasting disease pattern. (Table-5, Supplementary Table S1)
***8. PSP-Frontal (PSP-F):*** PSP-F (n=77; 7%) was notable for its non-motor onset profile, with 25% behavioral and 21% cognitive onset, far exceeding other subtypes. (Table-5) Family history of Parkinsonism was the highest (19%), raising the possibility of genetic or shared familial contributions. Mean age at onset was 62 years, with moderate disease duration (36 months). Diagnostic certainty was relatively high (91% “Probable PSP”). CGI severity often placed subjects in the moderate/mild illness spectrum, consistent with better functional preservation. Unlike PSP-RS or PSP-P, PSP-F showed more frequent diagnostic delay due to frontotemporal dementia-like presentations. This emphasizes PSP-F as a clinically challenging variant, often masquerading as psychiatric or behavioral syndromes.
***9. PSP-Cerebellar (PSP-C):*** Although PSP-C is not a formally recognized subtype in the MDS-PSP criteria, recruiting clinicians were permitted to classify phenotypes pragmatically based on the predominant presenting symptom or the most prominent symptom burden. Within this framework, PSP-C was documented as the “rarest” phenotype, likely reflecting limitations inherent to recruitment-based classification. PSP-C (n=13; 1.2%) cases had a mean onset age of 60 years, were uniformly motor-dominant at presentation (100%), and showed strong West Indian representation (54%). Diagnostic certainty was consistently high (100% “Probable PSP”). Disease duration was relatively long (48 months), and CGI severity ranged from moderate to severe, suggesting that while uncommon, PSP-C may follow a less aggressive course than PSP-RS or PSP-SL.

**Table 5 :** Major PSP Subtypes and their Features.

| **PSP Subtype** | **PSP-RS** | **PSP-P** | **PSP-CBS** | **PSP-PGF** | **PSP-PI** | **PSP-SL** | **PSP-OM** | **PSP-F** | **PSP-C** |
| --- | --- | --- | --- | --- | --- | --- | --- | --- | --- |
| **Demographic Features** |  |  |  |  |  |  |  |  |  |
| No. of Subjects | 420 | 186 | 119 | 54 | 72 | 14 | 23 | 77 | 13 |
| Male:Female<br>(Male, %) | 284:136<br>(67.6) | 127:59<br>(68.3) | 69:50<br>(58) | 38:16<br>(70.4) | 47:25<br>(65.3) | 13:1<br>(92.1) | 16:17<br>(48.5) | 52:25<br>(67.5) | 6:7<br>(46.2) |
| Mean age at<br>onset, years<br>(Range) | 61.6±7.<br>7 (41-85) | 62.9±8.0<br>(41-81) | 63.1±8.0<br>(38-83) | 63±8.3<br>(44-84) | 62±7.5<br>(41-77) | 67±7.3<br>(54-79) | 58.1±8.1<br>(44-77) | 62.4±8.7<br>(40-87) | 60.3±7.2<br>(48-75) |
| Duration of<br>symptoms,<br>months<br>Mean ±SD<br>(Range) | 38±25.4<br>(2-192) | 46.3±32.<br>8 (2-192) | 38.6±22<br>(4-108) | 35.8±21.<br>3 (6-96) | 35.9±23.<br>4 (6-144) | 25.9±13<br>(8-48) | 42.3±38.<br>1 (4-180) | 35.8±22.<br>8 (4-120) | 48.5±29.<br>9 (24-120) |
| Age at study<br>inclusion years<br>Mean ±SD<br>(Range) | 64.8±7.<br>8 (42-85) | 66.7±7.7<br>(50-83) | 66.3±8.0<br>(43-85) | 66±8.2<br>(49-88) | 65±7.7<br>(45-81) | 68.8±7.5<br>(55-81) | 61.4±8.9<br>(48-81) | 65.1±8.3<br>(42-83) | 64.1±6.5<br>(52-75) |
| Consanguineou | 40 (9.5) | 9 (5) | 6 (5) | 4 (7) | 4 (5) | 1 (7) | 0 (0) | 4 (5) | 0 (0) |
| s parentage (%) |  |  |  |  |  |  |  |  |  |
| Family history of Parkinsonism (%) | 18 (4.2) | 13 (7) | 6 (5) | 4 (7) | 5 (7) | 0 (0) | 2 (9) | 15 (19) | 0 (0) |
| Region of India: |  |  |  |  |  |  |  |  |  |
| 2. North | 79 (19) | 64 (34) | 31 (26) | 14 (26) | 25 (35) | 5 (36) | 6 (26) | 27 (35) | 3 (23) |
| 3. East | 116(28) | 19 (10) | 21 (18) | 4 (7) | 4 (6) | 0 (0) | 6 (26) | 10 (13) | 1 (8) |
| 4. West | 56 (13) | 51 (27) | 29 (24) | 18 (33) | 12 (17) | 3 (21) | 1 (4) | 28 (36) | 7 (54) |
| 5. South | 169 (40) | 52 (28) | 38 (32) | 18 (33) | 31 (43) | 6 (43) | 10 (43) | 12 (16) | 2 (15) |
| PSP-Rating score | 41±14.1<br>(1-90) | 33.9±13.<br>8 (1-74) | 42.6±19.<br>2 (1-85) | 32±13.1<br>(12-65) | 36.1±16.<br>4 (1-75) | 33.5±24.<br>1 (10-90) | 32±15.9<br>(11-82) | 35.5±16.<br>1 (2-84) | 34.7±13.<br>8 (18-67) |
| PSP-CDS | 10.1±3.<br>8 (1-21) | 9.0±3.9<br>(1-21) | 10.1±4.7<br>(1-20) | 8.7±3.8<br>(2-17) | 9.0±4.2<br>(1-19) | 9.1±4.7<br>(2-18) | 8.3±4.6<br>(1-20) | 9.1±4.1<br>(1-21) | 9.3±3.3<br>(5-16) |
| **Initial Symptom Category** |  |  |  |  |  |  |  |  |  |
| Behavioral (%) | 21 (5) | 5 (3) | 3 (3) | 0 (0) | 2 (3) | 0 (0) | 2 (9) | 19 (25) | 0 (0) |
| Cognitive (%) | 11 (3) | 4 (2) | 3 (3) | 0 (0) | 0 | 8 (57) | 0 (0) | 16 (21) | 0 (0) |
| Motor (%) | 386 (92) | 176 (95) | 112 (94) | 54 (100) | 70 (97) | 6 (43) | 21 (91) | 41 (53) | 13 (100) |
| **Diagnostic Certainty of PSP (MDS-PSP)** |  |  |  |  |  |  |  |  |  |
| Probably PSP (%) | 408 (97) | 167 (90) | 105 (88) | 45 (83) | 64 (89) | 12 (86) | 20 (87) | 70 (91) | 13 (100) |
| Possible PSP (%) | 10 (2) | 11 (6) | 8 (7) | 6 (11) | 6 (8) | 2 (14) | 3 (13) | 4 (5) | 0 (0) |
| Suggestive of PSP (%) | 2 (0.5) | 7 (4) | 6 (5) | 3 (6) | 2 (3) | 0 (0) | 0 (0) | 3 (4) | 0 (0) |

\*In 57 cases, primary phenotype was not specified in the records.

**Table 6:**
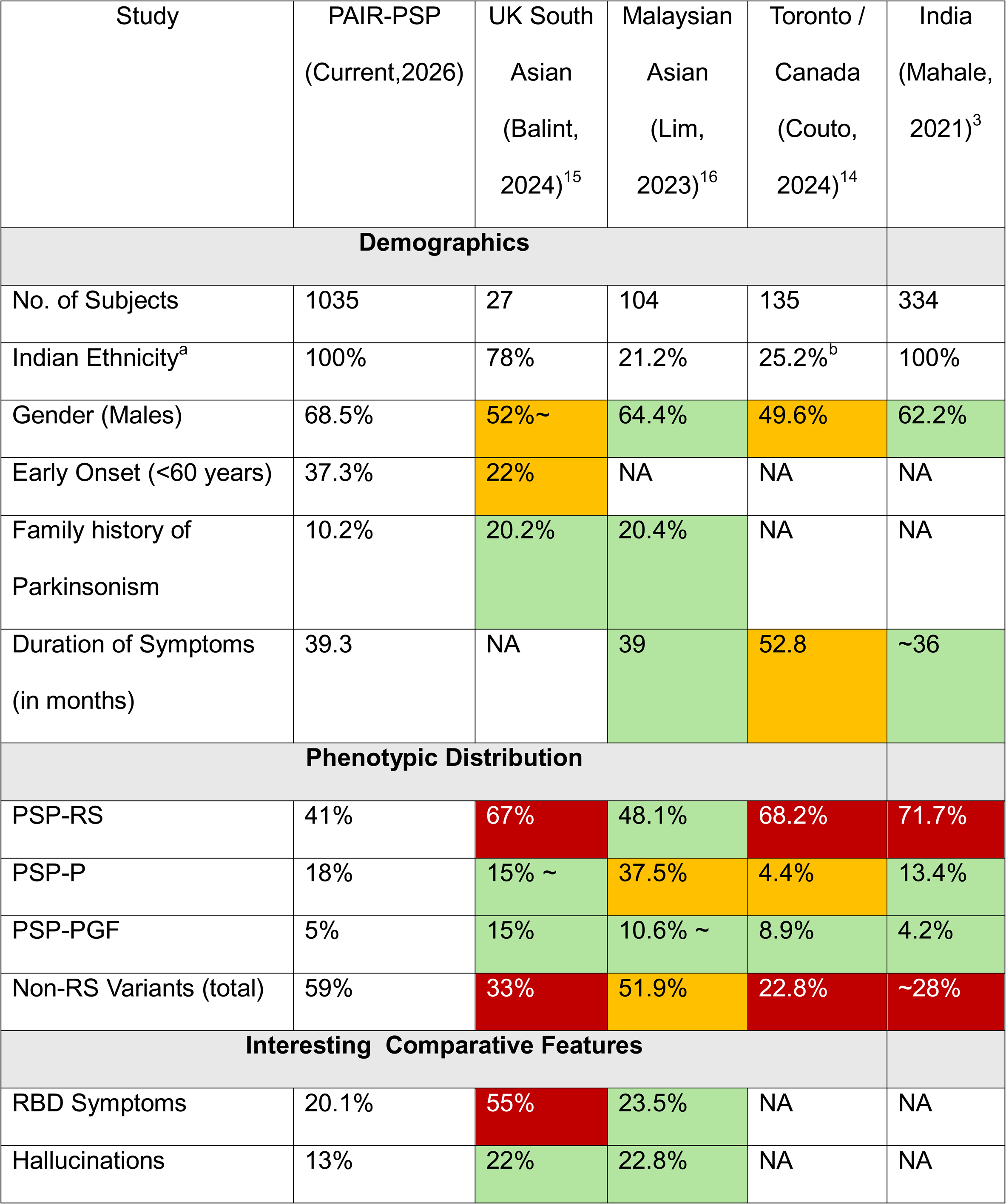
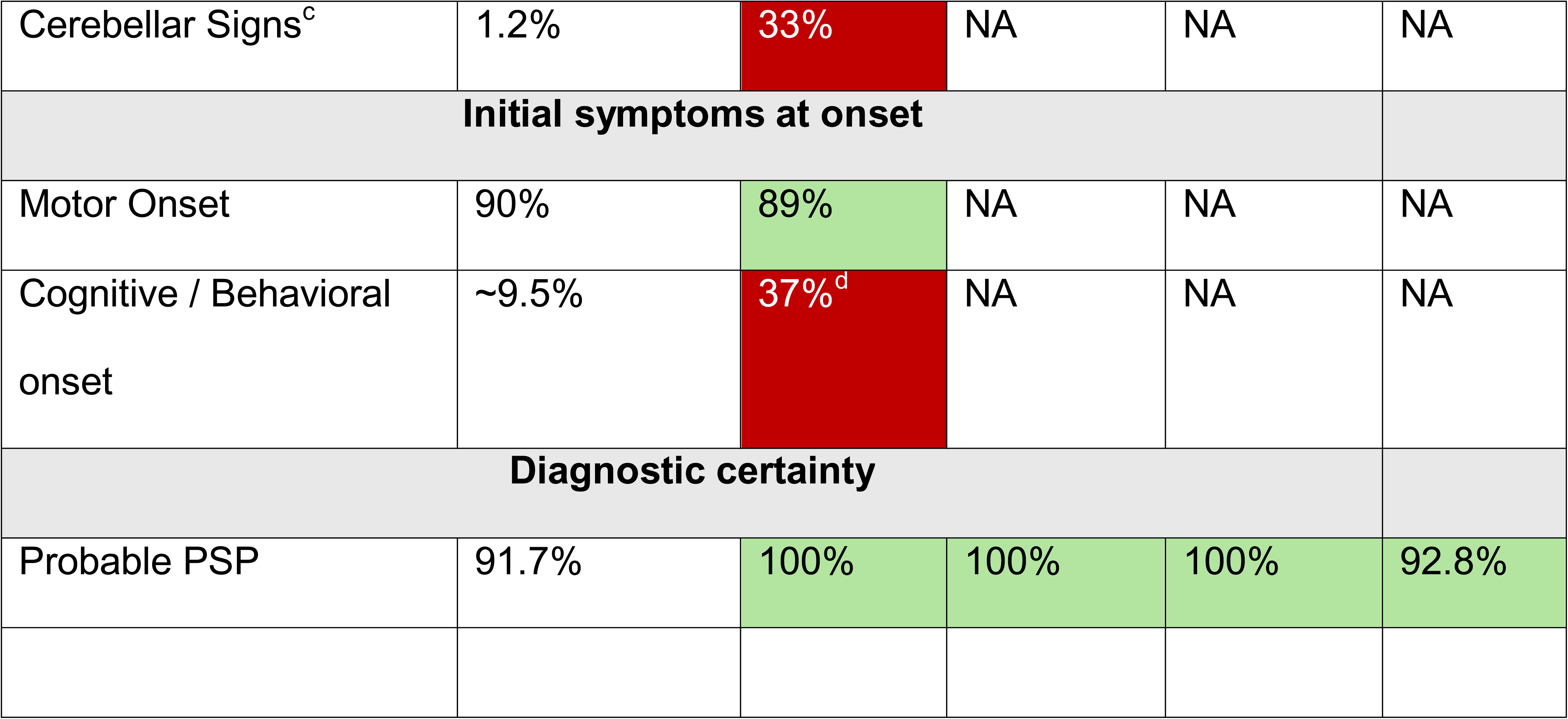
Comparison of Globally published Indian / South Asian Progressive Supranuclear Palsy Cohort Data with Current Study (PAIR-PSP Study)

| Study | PAIR-PSP<br>(Current,2026) | UK South<br>Asian<br>(Balint,<br>2024)<sup>15</sup> | Malaysian<br>Asian<br>(Lim,<br>2023)<sup>16</sup> | Toronto /<br>Canada<br>(Couto,<br>2024)<sup>14</sup> | India<br>(Mahale,<br>2021)<sup>3</sup> |
| --- | --- | --- | --- | --- | --- |
| **Demographics** |  |  |  |  |  |
| No. of Subjects | 1035 | 27 | 104 | 135 | 334 |
| Indian Ethnicity<sup>a</sup> | 100% | 78% | 21.2% | 25.2%<sup>b</sup> | 100% |
| Gender (Males) | 68.5% | 52%~ | 64.4% | 49.6% | 62.2% |
| Early Onset (<60 years) | 37.3% | 22% | NA | NA | NA |
| Family history of<br>Parkinsonism | 10.2% | 20.2% | 20.4% | NA | NA |
| Duration of Symptoms<br>(in months) | 39.3 | NA | 39 | 52.8 | ~36 |
| **Phenotypic Distribution** |  |  |  |  |  |
| PSP-RS | 41% | 67% | 48.1% | 68.2% | 71.7% |
| PSP-P | 18% | 15% ~ | 37.5% | 4.4% | 13.4% |
| PSP-PGF | 5% | 15% | 10.6% ~ | 8.9% | 4.2% |
| Non-RS Variants (total) | 59% | 33% | 51.9% | 22.8% | ~28% |
| **Interesting Comparative Features** |  |  |  |  |  |
| RBD Symptoms | 20.1% | 55% | 23.5% | NA | NA |
| Hallucinations | 13% | 22% | 22.8% | NA | NA |
| Cerebellar Signs<sup>c</sup> | 1.2% | 33% | NA | NA | NA |
| **Initial symptoms at onset** |  |  |  |  |  |
| Motor Onset | 90% | 89% | NA | NA | NA |
| Cognitive / Behavioral onset | ~9.5% | 37%<sup>d</sup> | NA | NA | NA |
| **Diagnostic certainty** |  |  |  |  |  |
| Probable PSP | 91.7% | 100% | 100% | 100% | 92.8% |

### Comparative Features of Subtypes

PSP-P clearly showed the strongest survival profile, increasing disproportionately in long-duration cohorts, while PSP-RS and PSP-SL declined. PSP-F and PSP-SL stood out with high rates of behavioral/cognitive onset, contrasting with the overwhelmingly motor-dominant onset of other subtypes. PSP-RS and PSP-C showed the highest rates of “Probable PSP,” whereas PSP-PGF and PSP-SL more frequently fell into the ‘Possible’ or ‘Suggestive of’ PSP categories, likely reflecting their broader differential diagnoses and the greater diagnostic uncertainty associated with these phenotypes.

PSP-PI and PSP-PGF showed clustering in South India, while PSP-F cases were widely distributed, with higher familial aggregation. Most subtypes retained male predominance, except PSP-OM (balanced) and PSP-SL (extreme male predominance, 92%).

### Treatment History and clinical history

Treatment patterns in the cohort were heterogeneous, though levodopa remained the most widely prescribed agent. A total of 893 subjects received levodopa, with mean daily dosages ranging from 300 to 400 mg. Subjective improvement was documented in 544 patients, though sustained benefit was limited: only 186 patients reported >25% perceived improvement, while 358 experienced <25% benefit. Amantadine was the second most frequently used medication (n=351), with 177 patients reporting some degree of improvement. Various other medications, pharmacological and non-pharmacological therapies, including alternative therapies, were also tried. Across the cohort, the disease burden was substantial. The mean PSP Rating Scale (PSPRS) score was 37.9 ± 15.6 (range 1–90), reflecting moderate to advanced disease severity at evaluation. Similarly, the mean PSP Clinical Deficits Scale (PSP-CDS) was 9.5 ± 4.1 (range 1–21), further corroborating the high functional impairment in daily activities.

These findings, in conjunction with milestone analyses, highlight that despite symptomatic therapies, most patients progress to significant disability within a relatively short timeframe, reinforcing the urgent need for effective disease-modifying strategies.

### Evolution of Clinical Milestones Across PSP Subtypes

Milestone analysis(9) highlights distinct trajectories across PSP subtypes, but is limited by sparse data. (Supplementary Table- S1) Falls (>2/year) emerged earliest in PSP-PGF (mean 13.7 months) and PSP-SL (16.3 months), while PSP-P developed falls later (31.2 months), consistent with its slower progression. Cognitive disability appeared most rapidly in PSP-PI (13 months) and PSP-SL (15 months), contrasting with PSP-RS (26 months) and PSP-P (24 months). Urinary milestones showed marked subtype variability: PSP-P developed late urinary dependence (50.7 months), whereas PSP-RS and PSP-CBS reached this earlier (∼27–36 months). Severe dysphagia was relatively infrequent but occurred earliest in PSP-P (24 months), while PSP-F showed the longest latency (44 months). Speech declined fastest in the PSP-SL subtype (reaching unintelligibility in 19.8 months), while CBS, OM, and PGF progressed about twice as slowly (∼40 months).–Loss of independent mobility clustered between 27–35 months in most groups, but PSP-P again showed delayed progression (34.8 months).

Clinical severity ratings reinforced these patterns: PSP-RS patients were predominantly “markedly to moderately ill,” while PSP-P and PSP-F included more in the “moderate/mild” range, aligning with their relative survival advantage. Thus, milestone evolution underscores the aggressive course of PSP-RS and PSP-SL, contrasted with the slower progression of PSP-P and PSP-F, offering important insights into patient care and prognosis.

## Discussion

Progressive supranuclear palsy is a rare yet devastating neurodegenerative tauopathy with profound impact on patients and caregivers. Despite being described six decades ago, global data on PSP remain limited, with most clinical, pathological, and genetic insights derived from Europe and North America.(10,11) Emerging evidence highlights considerable regional variation in PSP prevalence, clinical phenotypes, and prognosis, shaped by both genetic predispositions (e.g., MAPT haplotype differences) and environmental exposures. In this context, data from India are particularly valuable, given its large population, high genetic diversity, and distinctive environmental and healthcare contexts.(2,6) The paucity of community-based and multicentric data from India / Asia has long been a barrier to global PSP research.(12) Our study, therefore, represents an important step toward bridging this gap, adding much-needed representation from South Asia to the global PSP picture.

Our pan-India multicenter cohort provides the most extensive systematic clinical profiling of PSP from this region to date. This cohort of 1035 would be one of the largest global series on clinical profile of PSP and add considerably to the under represented population data. The mean age at onset (62.4 years) in our cohort is slightly younger than that reported in European and North American series (typically 63–72 years), aligning with earlier observations of earlier onset in South Asians. The male predominance (68.5%) observed in this cohort warrants cautious interpretation. While similar sex distribution has been reported in prior Indian PSP studies, the present dataset was not designed to distinguish between biological susceptibility, referral patterns, or sociocultural influences on healthcare access and case ascertainment.

These factors merit dedicated evaluation in future studies. Few studies globally including from Canada(13,14), United Kingdom(15) and Malaysia(16) have tried to specifically highlight on the south Asian clusters, broadly agree on early age of onset and higher family history burden. (Table-6) They do diverge on phenotypic distribution and atypical features, which can be largely explained based upon data collection protocols rather than necessarily on true biological differences between the cohorts.

PSP-RS was the predominant subtype, but the proportions of PSP-P and PSP-CBS were higher than in some Western cohorts, echoing previous Indian single-center studies.(3,6,17–20) Importantly, early postural instability and gaze palsy remained dominant across subtypes, but phenotypic heterogeneity was pronounced, with behavioral and cognitive onset being notable in subsets of younger patients. The shift from ‘Probable PSP’ (94%) toward ‘Possible PSP’(86%) with later age at onset may indicate that older individuals more often present with less typical or diagnostically less specific syndromes. While alternative diagnoses were considered during standard clinical work-up, the present dataset was not designed to formally distinguish the relative contribution of phenotypic subtlety, disease evolution, or overlap with age-related mimics. In current study family history data were derived from routine clinical documentation and were primarily focused on parkinsonism and dementia; broader psychiatric and neurobehavioral family histories were not uniformly captured across centers. This may have led to underestimation of familial aggregation in certain phenotypes, particularly PSP-F, where overlap with cognitive and frontotemporal syndromes may be relevant. (Table-6) Our longitudinal milestone analyses demonstrated that PSP-RS had the most rapid evolution to falls, dysphagia, and wheelchair dependence, while PSP-P showed a relatively protracted course — mirroring global natural history reports.(21–25) PSP-C remains a challenging subtype within the MDS-PSP framework, as cerebellar features are not explicitly weighted in the current criteria. Patients may initially present with cerebellar symptoms and later evolve into a classical PSP phenotype, highlighting limitations of recruitment-based classification and variability in its reported rarity. Further studies are needed to clarify whether these cases represent true PSP/tauopathy or alternative etiologies, and to determine if current diagnostic criteria require refinement. Collectively, these findings reinforce both the universality of PSP’s core clinical burden and the importance of regional differences in phenotype frequency, age of onset, and treatment utilization.

A major strength lies in its multicenter design under the PRAI, which ensured standardized recruitment, specialist-driven diagnosis (using MDS-PSP criteria), and centralized data capture. The collaboration with MedGenome and integration with the Global Parkinson’s Genetic Program adds further robustness and genetic translational potential. This is the first pan-India effort to capture detailed demographic, clinical, and treatment variables alongside genetic sampling, thereby setting a benchmark for future work. The large sample size across diverse Indian states enhances generalizability, especially compared to prior single-center reports.

Several limitations merit acknowledgement. First, clinical phenotypes of PSP can evolve with disease duration — patients initially meeting criteria for PSP-P, PSP-PGF, or other subtypes may ultimately evolve into PSP-RS. This temporal evolution complicates phenotype assignment and introduces heterogeneity, a challenge also emphasized in global literature. Within the speech/language subgroup, documentation did not uniformly distinguish progressive apraxia of speech from nonfluent/agrammatic language impairment, and classification was therefore based on clinical assignment to the MDS-PSP C1 domain rather than a standardized formal speech-language battery. Second, while multicentric, the cohort still reflects referral center populations, introducing a possible bias toward more severe or atypical cases. Third, longitudinal follow-up data remain limited; survival analyses and imaging/biomarker correlations will strengthen future outputs. Finally, although genetic samples have been collected, analyses are ongoing; integration of genotype-phenotype correlations will be critical to resolve whether MAPT haplotype and other genetic modifiers contribute to the regional differences observed.

This study underscores both the commonalities and unique features of PSP in India. Future efforts must focus on harmonized prospective follow-up, biomarker integration, and genetic analyses within global consortia to delineate ancestry-specific risk factors and clinical trajectories. Collaborative approaches will be essential to define how phenotype evolution relates to underlying pathology and to build predictive models for progression. Ultimately, insights from large, diverse populations such as India will be indispensable in informing equitable global clinical trials and advancing the search for disease-modifying therapies.

## Data Availability

Data used in the preparation of this article were obtained from the Global Parkinsons Genetics Program (GP2; https://gp2.org). GP2 data can be requested through AMP PD (https://amp-pd.org). Data used in this manuscript will be shared through GP2, specifically in Tier 2. Data will be available in GP2 release 13 (https://doi.org/10.5281/zenodo.7904831). Data was generated in India as a part of a collaboration between Parkinsons Research Alliance of India (PRAI), GP2, MedGenome India, and KS Bio Clinserv. The data is located in a cloud server that is hosted in India. Data cannot be downloaded and may only be accessed by approved and verified researchers.

## Acknowledgments

This project was supported by the Global Parkinson’s Genetics Program (GP2; https://gp2.org). GP2 is funded by the Aligning Science Across Parkinson’s (ASAP) (https://ror.org/03zj4c476) initiative and implemented by The Michael J. Fox Foundation for Parkinson’s Research (MJFF) (https://ror.org/03arq3225). For a complete list of GP2 members see https://doi.org/10.5281/zenodo.7904831.

## Data Availability

Data used in the preparation of this article were obtained from the Global Parkinson’s Genetics Program (GP2; https://gp2.org). GP2 data can be requested through AMP PD (https://amp-pd.org). Data used in this manuscript will be shared through GP2, specifically in Tier 2. Data will be available in GP2 release 13 (https://zenodo.org/records/17753486). Data was generated in India as a part of a collaboration between Parkinson’s Research Alliance of India (PRAI), GP2, MedGenome India, and KS Bio Clinserv. The data is located in a cloud server that is hosted in India. Data cannot be downloaded and may only be accessed by approved and verified researchers.

## Code Availability

No new code is generated in relation to this manuscript. The images were generated for the study from the core clinical data and using design platforms of Canva and its plugins

## Ethical Publication Guidelines of the Journal

All participating centers obtained independent Institutional Ethics Committee/Review Board approvals for prospective patient recruitment from their respective ethics / Review boards.

1. The project was reviewed by respective clinical recruitment centers reviewed / ethics boards and approval for prospective patient recruitment from their center was obtained.
2. All subjects reviewed the written approved patient information sheet and patient consent forms from the review boards. The consents forms were explained by respective centers investigator / research associate in a language understandable by the patient / caregiver. The consent forms were available in English and local dialects of respective geographic territory.
3. All authors have read and compiled with the Journal’s ethical publication guidelines – “We confirm that we have read the Journal’s position on issues involved in ethical publication and affirm that this work is consistent with those guidelines”.

## Author Roles

1. Research project: A. Conception, B. Organization, C. Execution;
2. Statistical Analysis: A. Design, B. Execution, C. Review and Critique;
3. Manuscript Preparation: A. Writing of the first draft, B. Review and Critique;

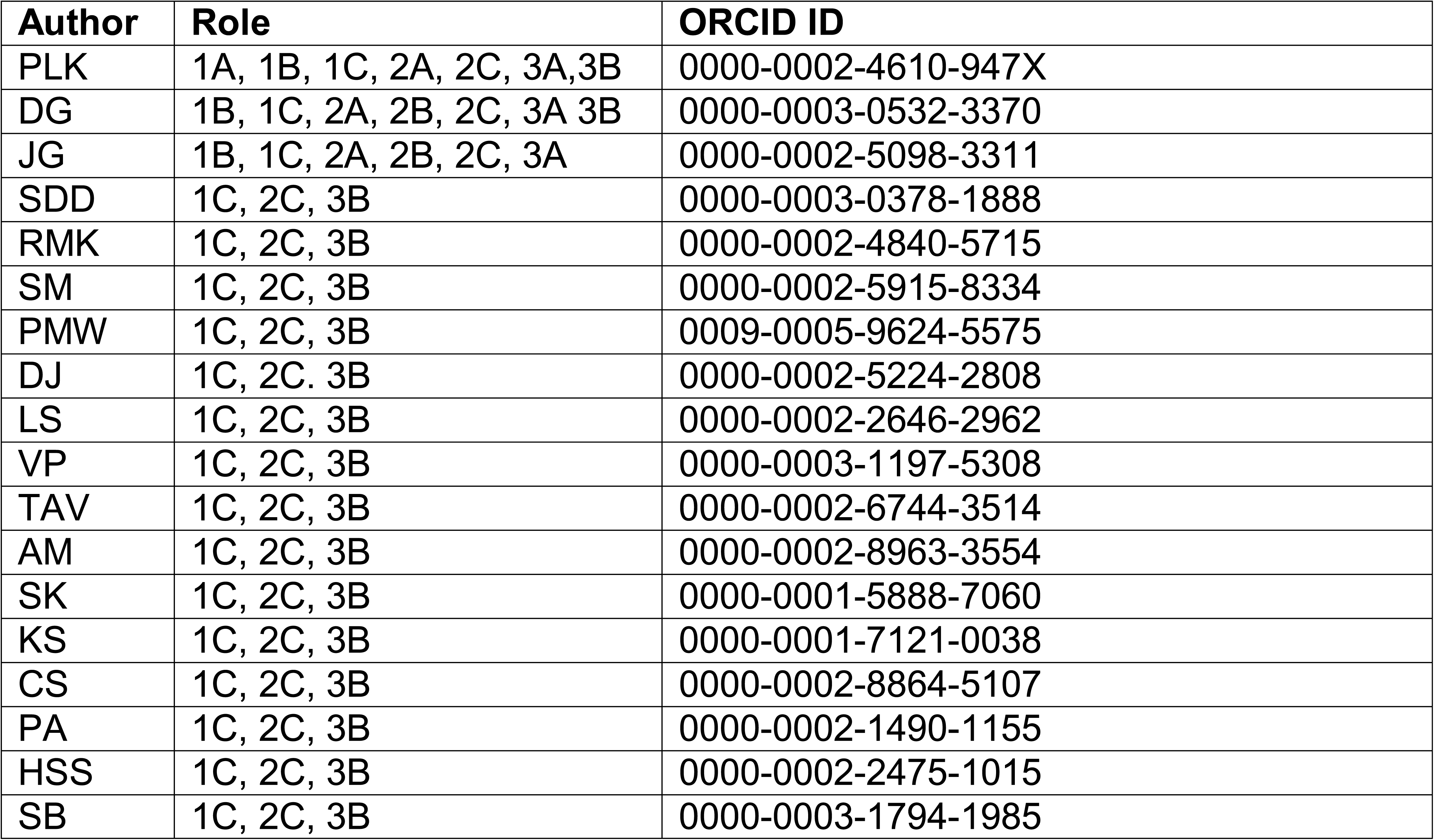
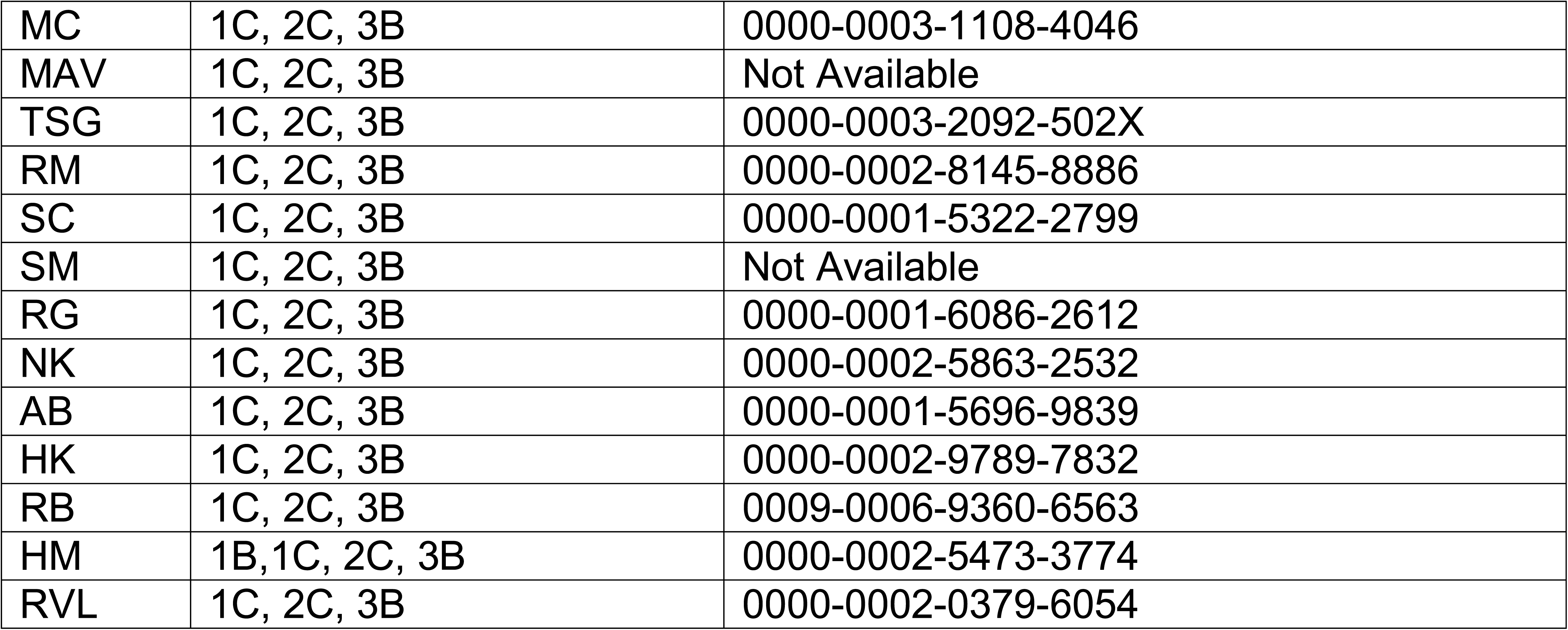

## Funding sources for study

Global Parkinson’s Genetic Program (GP2)

## Financial Disclosures

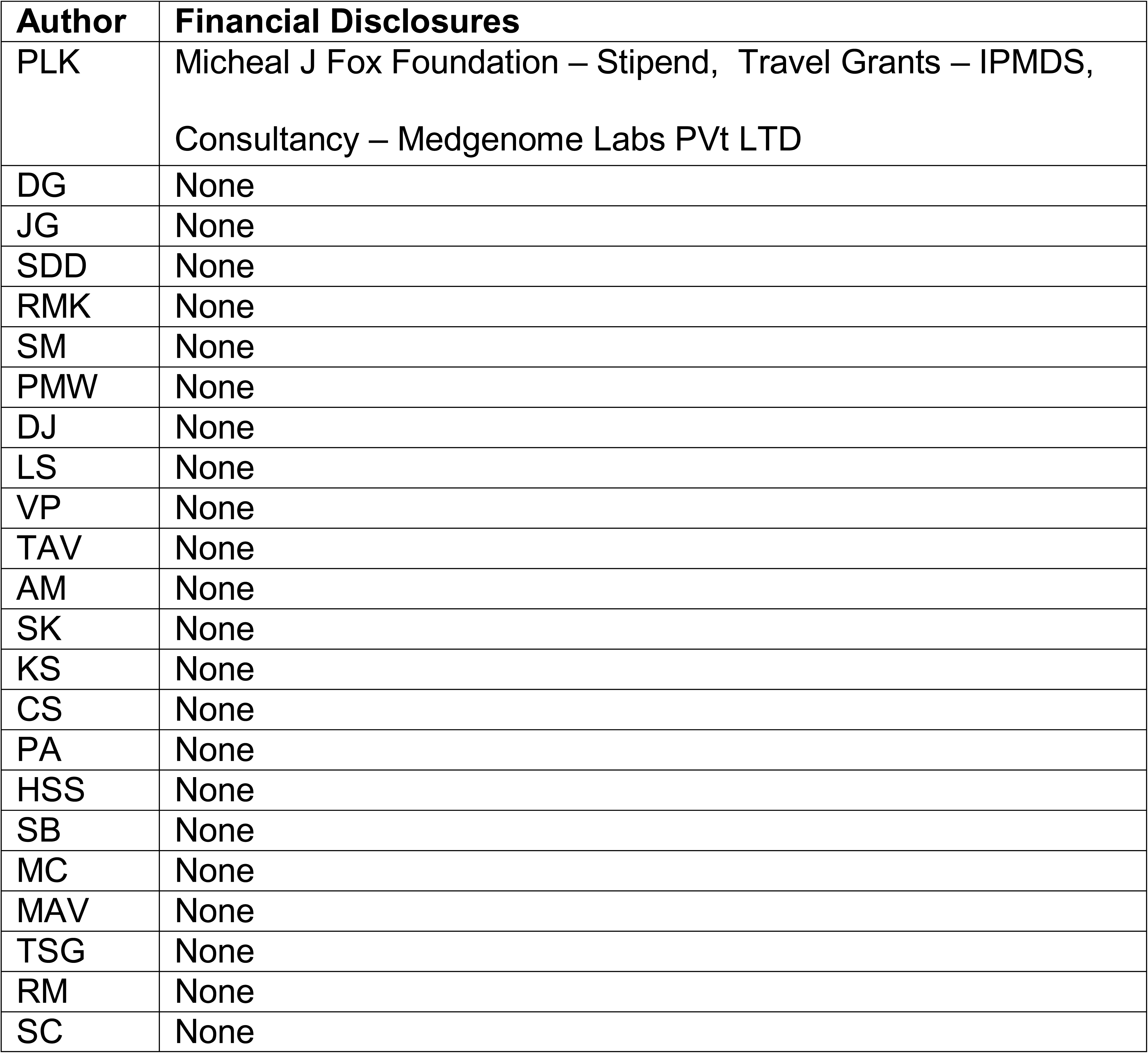
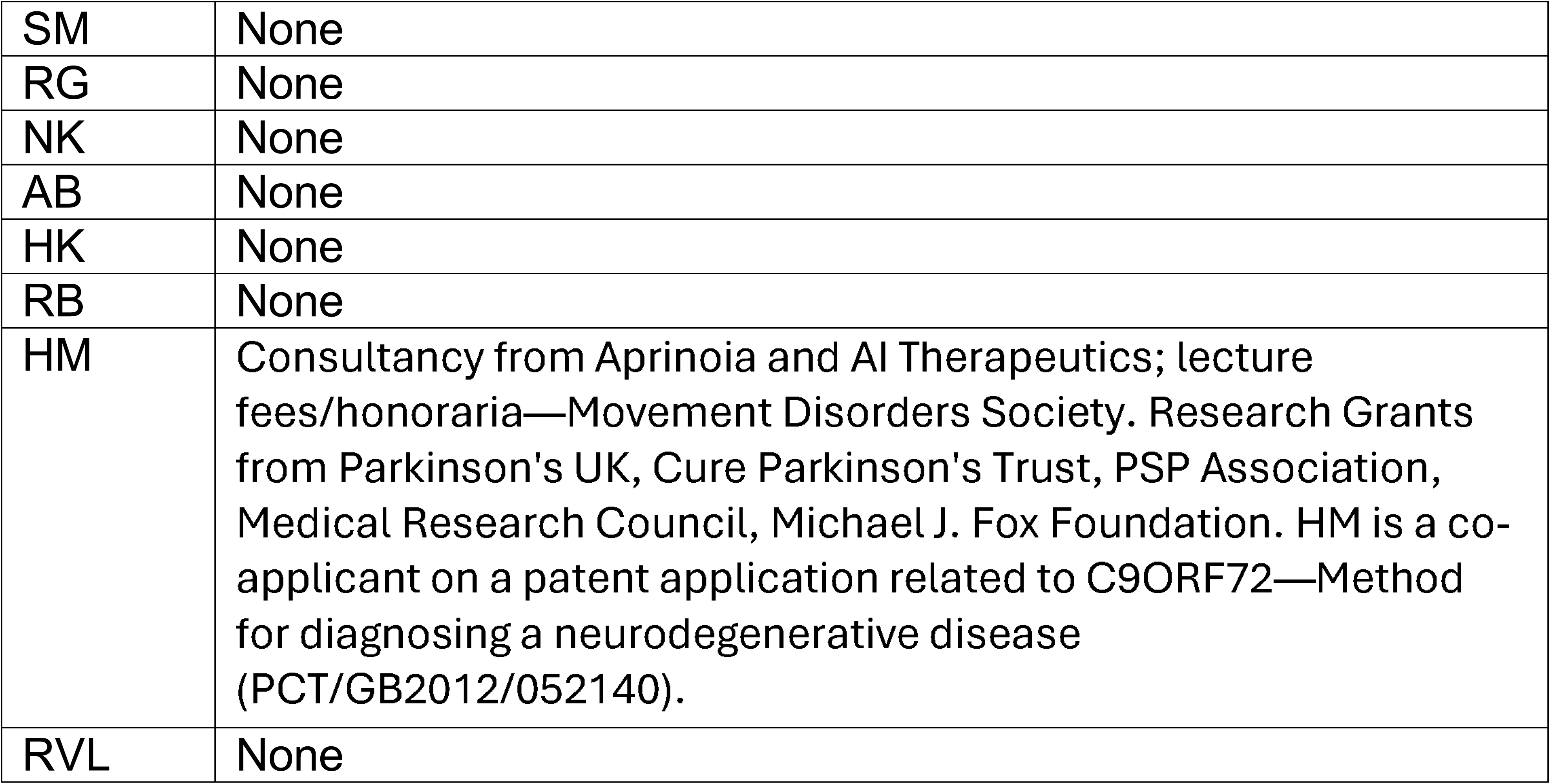

| **Author** | **Financial Disclosures** |
| --- | --- |
| PLK | Micheal J Fox Foundation – Stipend, Travel Grants – IPMDS,<br>Consultancy – Medgenome Labs PVt LTD |
| DG | None |
| JG | None |
| SDD | None |
| RMK | None |
| SM | None |
| PMW | None |
| DJ | None |
| LS | None |
| VP | None |
| TAV | None |
| AM | None |
| SK | None |
| KS | None |
| CS | None |
| PA | None |
| HSS | None |
| SB | None |
| MC | None |
| MAV | None |
| TSG | None |
| RM | None |
| SC | None |
| SM | None |
| RG | None |
| NK | None |
| AB | None |
| HK | None |
| RB | None |
| HM | Consultancy from Aprinoia and AI Therapeutics; lecture fees/honoraria—Movement Disorders Society. Research Grants from Parkinson's UK, Cure Parkinson's Trust, PSP Association, Medical Research Council, Michael J. Fox Foundation. HM is a co-applicant on a patent application related to C9ORF72—Method for diagnosing a neurodegenerative disease (PCT/GB2012/052140). |
| RVL | None |

## Funding Sources and Conflict of Interest

The authors declare that there are no conflicts of interest relevant to this work.

Supplementary Table - S1 : Clinical Milestones Evolution

Supplementary Figure A: Initial Presenting Symptoms at onset

Supplementary Figure B: Documented Motor and Occulomotor Symptoms till the recruitment The list shows symptoms noted during the course of symptoms and may not indicate as presenting symptom or the symptom at the point of inclusion The list indicates the symptoms were documented. Undocumented indicates either absence of symptoms or as data not entered

Supplementary Figure C: Documented Non Motor symptoms during the course The list shows symptoms noted during the course of symptoms and may not indicate as presenting symptom or the symptom at the point of inclusion The list indicates the symptoms were documented. Undocumented indicates either absence of symptoms or as data not entered

##Appendix -1 : Study Conduct and Methodological Framework of the PAIR-PSP Project

## Notes

### Competing Interest Statement

The authors have declared no competing interest.

### Funding Statement

This project was supported by the Global Parkinsons Genetics Program (GP2; https://gp2.org). GP2 is funded by the Aligning Science Across Parkinsons (ASAP) (https://ror.org/03zj4c476) initiative and implemented by The Michael J. Fox Foundation for Parkinsons Research (MJFF) (https://ror.org/03arq3225). For a complete list of GP2 members see https://doi.org/10.5281/zenodo.7904831.

### Author Declarations

Ethics Committe / IRB of Rajarajeswari Medical College and Hospital gave ethical approval for this work (IEC Approval No. RRMCH-IEC/11/2024.). Further being an multicenter study, each center have got their independent ethics board approvals

### Summary of Updates

This is an updated peer reivewed version accepted for publication in MDCP. Has extra table in main manuscript and new supplementary figures showing motor and non motor features.

## References

1. Prashanth L, Kumar H, Wadia P, Muthane U. The spectrum of movement disorders in tertiary care centers in India: A tale of three cities. Ann Indian Acad Neurol. 2021;24(5):721–5. doi:10.4103/aian.AIAN_1257_20

2. Raju S, Shetty K, Sahoo L, Paramanandam V, Iyer JM, Bowmick S, et al. Progressive Supranuclear Palsy in India: Past, Present, and Future. Annals of Indian Academy of Neurology. 2025 Jan;28(1):17–25. doi:10.4103/aian.aian_515_24

3. Mahale R, Krishnan S, Divya K, Jisha V, Kishore A. Subtypes of PSP and prognosis: A retrospective analysis. Ann Indian Acad Neurol. 2021;24(1):56. doi:10.4103/aian.AIAN_611_20

4. Chaithra SP, Prasad S, Holla VV, Stezin A, Kamble N, Yadav R, et al. The Non-Motor Symptom Profile of Progressive Supranuclear Palsy. JMD. 2020 May 31;13(2):118–26. doi:10.14802/jmd.19066

5. Batla A, Nehru R, Vijay T. Vertical wrinkling of the forehead or Procerus sign in Progressive Supranuclear Palsy. Journal of the Neurological Sciences. 2010 Nov;298(1–2):148–9. doi:10.1016/j.jns.2010.08.010

6. Kukkle PL, Neupane R, Pantelyat A, Wills A, Jabbari E, Dopper EGP, et al. Progressive Supranuclear Palsy—A Global Review. Movement Disord Clin Pract. 2025 Sep 3;mdc3.70338. doi:10.1002/mdc3.70338

7. Singleton A, Blauwendraat C, Morris HR, Noyce AJ, Klein C, Mata I, et al. Parkinson’s disease: emerging opportunities through global collaboration. Lancet. 2026 Jan 17;407(10525):203–5. doi:10.1016/S0140-6736(25)01910-5 PubMed PMID: 41077048.

8. Höglinger GU, Respondek G, Stamelou M, Kurz C, Josephs KA, Lang AE, et al. Clinical diagnosis of progressive supranuclear palsy: The movement disorder society criteria: MDS Clinical Diagnostic Criteria for PSP. Mov Disord. 2017 Jun;32(6):853–64. doi:10.1002/mds.26987

9. O’Sullivan SS, Massey LA, Williams DR, Silveira-Moriyama L, Kempster PA, Holton JL, et al. Clinical outcomes of progressive supranuclear palsy and multiple system atrophy. Brain. 2008 May;131(5):5. doi:10.1093/brain/awn065

10. Steele JC. Progressive Supranuclear Palsy: A Heterogeneous Degeneration Involving the Brain Stem, Basal Ganglia and Cerebellum With Vertical Gaze and Pseudobulbar Palsy, Nuchal Dystonia and Dementia. Arch Neurol. 1964 Apr 1;10(4):333. doi:10.1001/archneur.1964.00460160003001

11. Boxer AL, Yu JT, Golbe LI, Litvan I, Lang AE, Höglinger GU. Advances in progressive supranuclear palsy: new diagnostic criteria, biomarkers, and therapeutic approaches. Lancet Neurol. 2017 Jul;16(7):552–63. doi:10.1016/S1474-4422(17)30157-6 PubMed PMID: 28653647; PubMed Central PMCID: PMC5802400.

12. Shah H, Kukkle PL. Barriers to providing movement disorders care in India. Current Opinion in Neurology. 2025 Aug;38(4):361–9. doi:10.1097/WCO.0000000000001375

13. Couto B, Di Luca DG, Antwi J, Bhakta P, Fox S, Tartaglia MC, et al. Ethnic background and distribution of clinical phenotypes in patients with probable progressive supranuclear palsy. Parkinsonism & Related Disorders. 2024 Jun;123:106955. doi:10.1016/j.parkreldis.2024.106955

14. Couto B, Tartaglia MC, Kovacs GG, Lang AE. Differences in progressive supranuclear palsy in patients of Asian ancestry? Parkinsonism & Related Disorders. 2025 Jan;130:107179. doi:10.1016/j.parkreldis.2024.107179

15. Balint B, Neo S, Magrinelli F, Mulroy E, Latorre A, Stamelou M, et al. Ethnic Differences in Atypical Parkinsonism-is South Asian PSP Different? Mov Disord Clin Pract. 2024 Nov;11(11):1355–64. doi:10.1002/mdc3.14182 PubMed PMID: 39113437; PubMed Central PMCID: PMC11542300.

16. Lim SY, Dy Closas AMF, Tan AH, Lim JL, Tan YJ, Vijayanathan Y, et al. New insights from a multi-ethnic Asian progressive supranuclear palsy cohort. Parkinsonism Relat Disord. 2023 Mar;108:105296. doi:10.1016/j.parkreldis.2023.105296 PubMed PMID: 36682278.

17. Fujioka S, Algom AA, Murray ME, Sanchez-Contreras MY, Tacik P, Tsuboi Y, et al. Tremor in progressive supranuclear palsy. Parkinsonism & Related Disorders. 2016 Jun;27:93–7. doi:10.1016/j.parkreldis.2016.03.015

18. Hewer S, Varley S, Boxer AL, Paul E, Williams DR, on behalf of the AL_-_108_-_231 Investigators. Minimal clinically important worsening on the progressive supranuclear Palsy Rating Scale. Movement Disorders. 2016 Oct;31(10):10. doi:10.1002/mds.26694

19. Nasri A, Sghaier I, Neji A, Gharbi A, Abida Y, Mrabet S, et al. Phenotypic Spectrum of Progressive Supranuclear Palsy: Clinical Study and Apolipoprotein E Effect. JMD. 2024 Apr 30;17(2):2. doi:10.14802/jmd.23178

20. Painous C, Fernández M, Cámara A, Alba_-_Arbalat S, Soto M, Brenlla C, et al. Barcelona Progressive Supranuclear Palsy Registry: Clinical, Oculomotor, and Cerebrospinal Fluid Markers; from Suggestive to Definite Cases. Movement Disorders. 2025 Oct 29;mds.70086. doi:10.1002/mds.70086

21. Litvan I, Mangone CA, McKee A, Verny M, Parsa A, Jellinger K, et al. Natural history of progressive supranuclear palsy (Steele-Richardson-Olszewski syndrome) and clinical predictors of survival: a clinicopathological study. Journal of Neurology, Neurosurgery & Psychiatry. 1996 Jun 1;60(6):6. doi:10.1136/jnnp.60.6.615

22. Golbe LI, Davis PH, Schoenberg BS, Duvoisin RC. Prevalence and natural history of progressive supranuclear palsy. Neurology. 1988 Jul;38(7):1031–4. doi:10.1212/wnl.38.7.1031 PubMed PMID: 3386818.

23. Nath U, Ben-Shlomo Y, Thomson RG, Lees AJ, Burn DJ. Clinical features and natural history of progressive supranuclear palsy: A clinical cohort study. Neurology. 2003 Mar 25;60(6):6. doi:10.1212/01.WNL.0000052991.70149.68

24. Papapetropoulos S, Gonzalez J, Mash DC. Natural history of progressive supranuclear palsy: a clinicopathologic study from a population of brain donors. Eur Neurol. 2005;54(1):1–9. doi:10.1159/000086754 PubMed PMID: 16015014.

25. Respondek G, Stamelou M, Kurz C, Ferguson LW, Rajput A, Chiu WZ, et al. The phenotypic spectrum of progressive supranuclear palsy: A retrospective multicenter study of 100 definite cases. Movement Disorders. 2014 Dec;29(14):14. doi:10.1002/mds.26054

