## Supplementary figures and images for "Progressive Supranuclear Palsy in India: Insights from a Large Multicenter Clinical Cohort (Project PAIR-PSP)"

### Supplementary Figure B

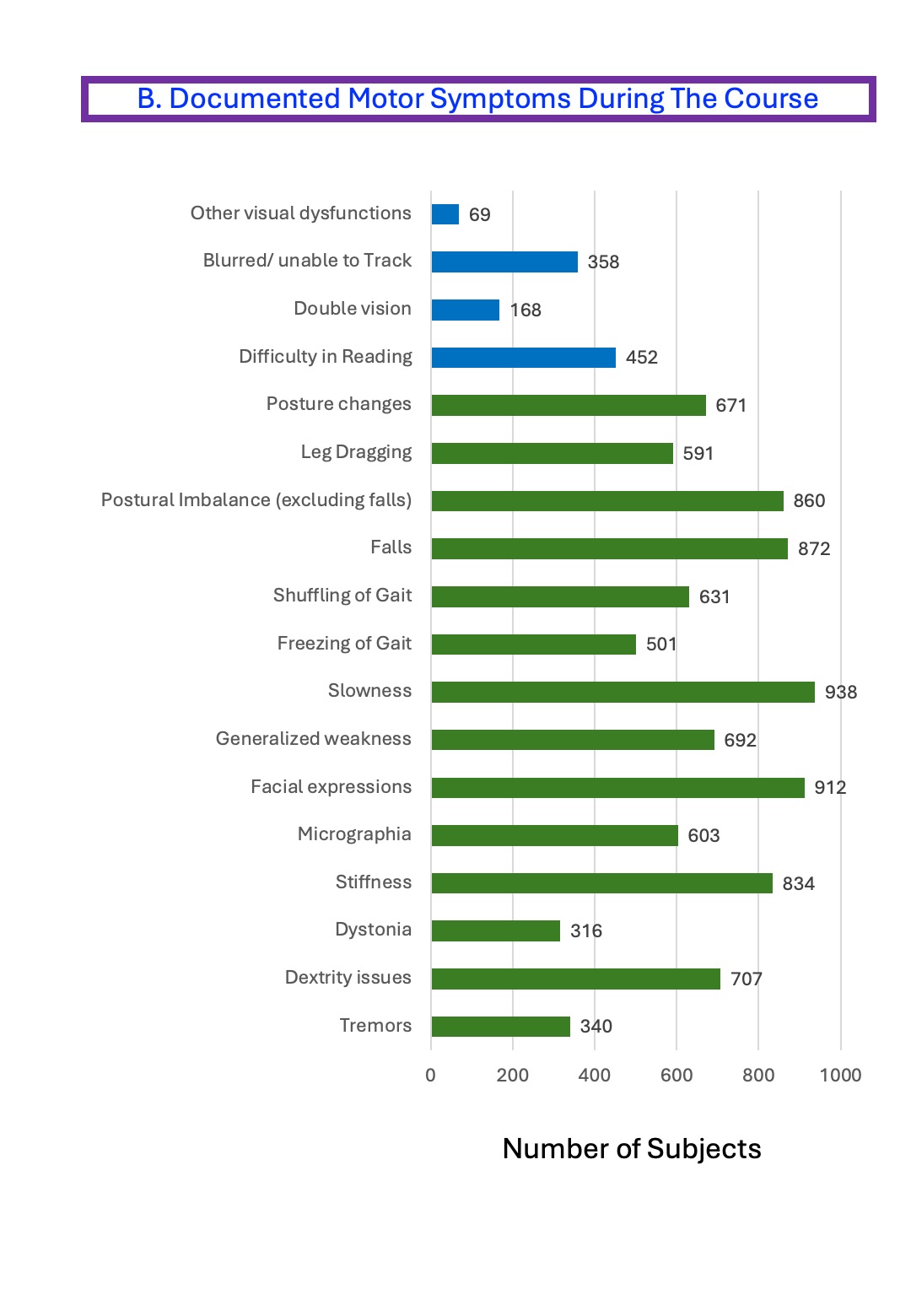

### Supplementary Figure C

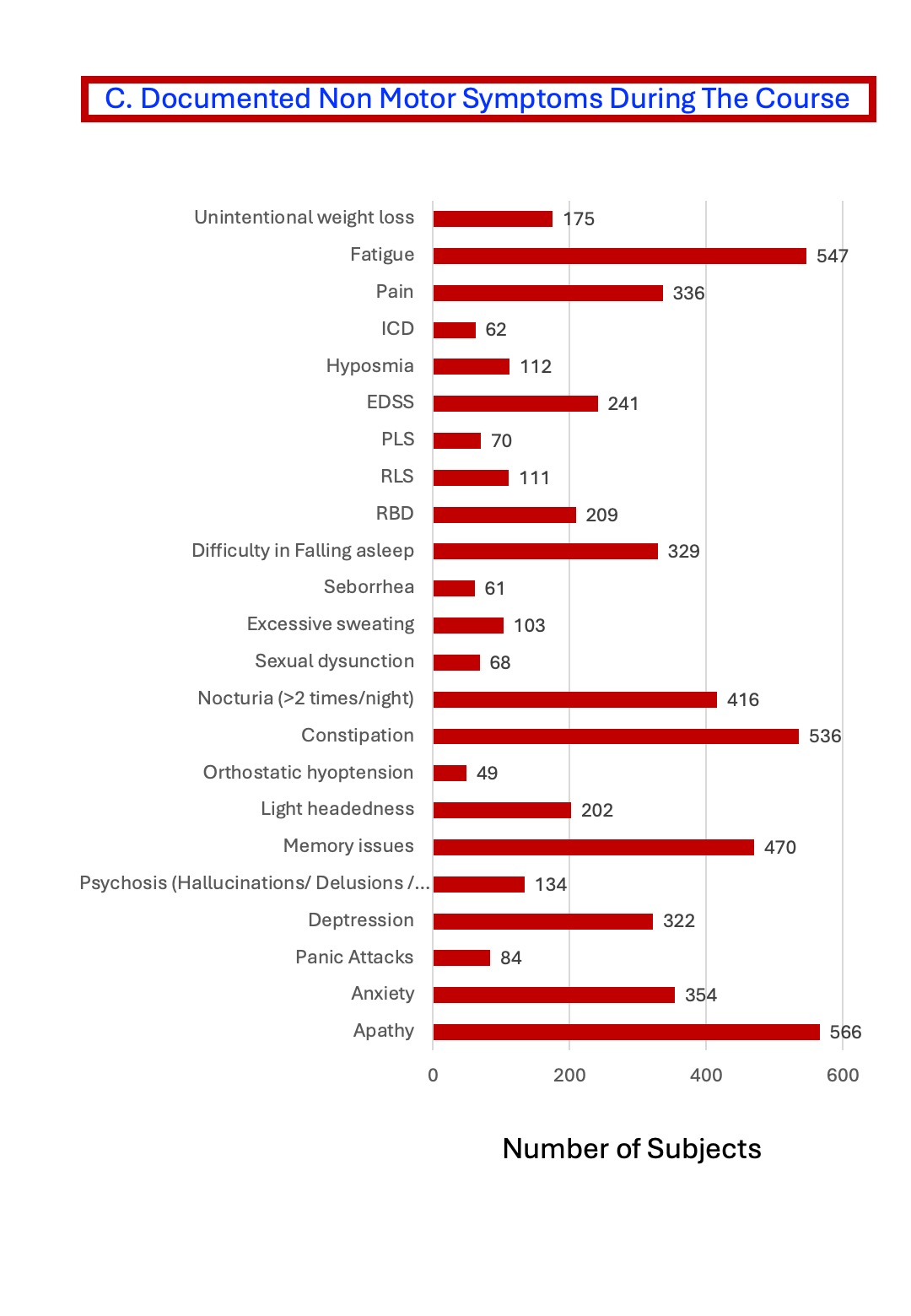

### Supplementary Figure-A

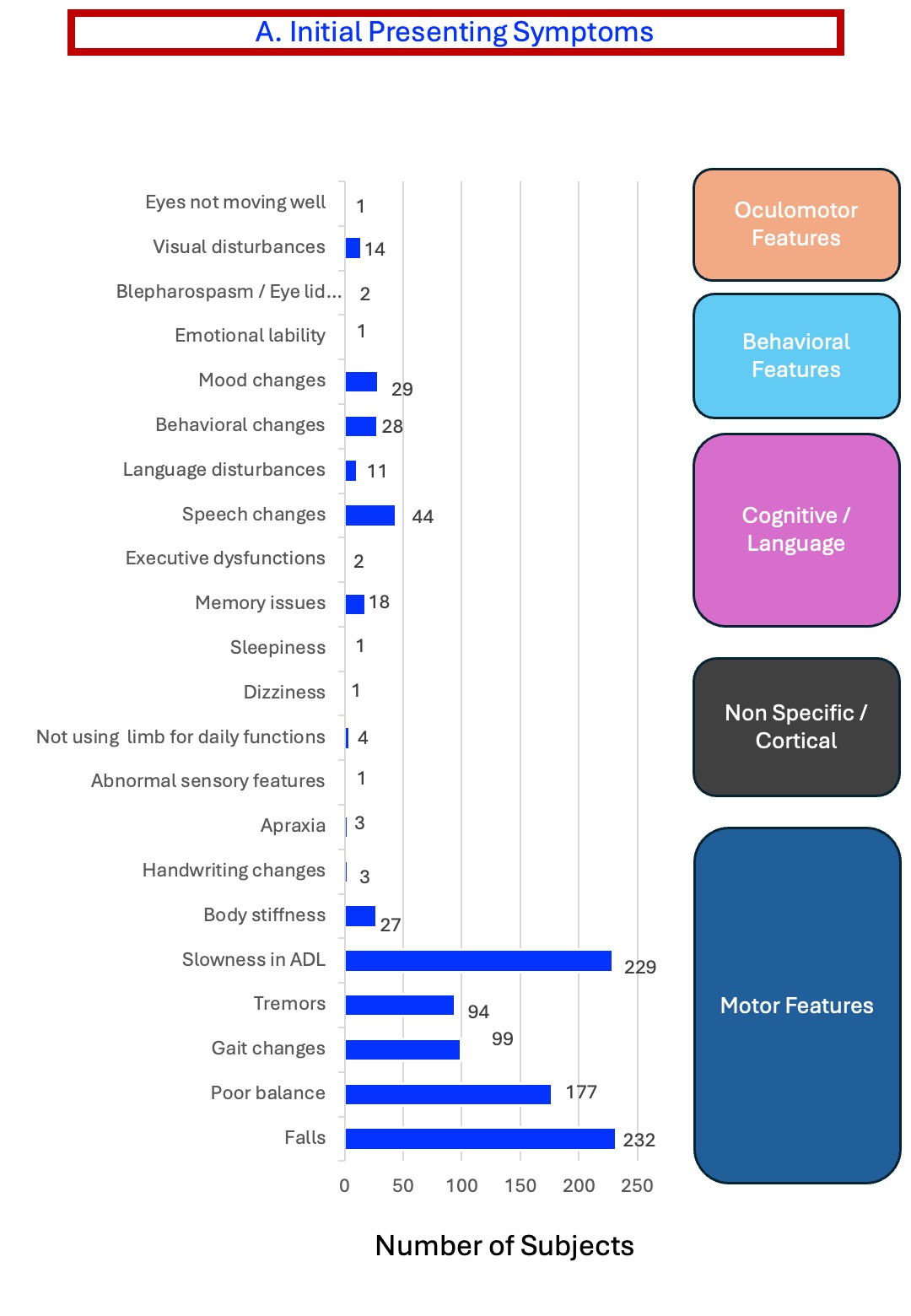
