## Supplementary Table for "Progressive Supranuclear Palsy in India: Insights from a Large Multicenter Clinical Cohort (Project PAIR-PSP)"

**Supplementary Table - S1 : Clinical Milestones Evolution:**

| **PSP Subtype** | **PSP-RS** | **PSP-P** | **PSP-CBS** | **PSP-PGF** | **PSP-PI** | **PSP-SL** | **PSP-OM** | **PSP-F** | **PSP-C** |
| --- | --- | --- | --- | --- | --- | --- | --- | --- | --- |
| **Demographic Features** | | | | | | | | | |
| No. of Subjects | 420 | 186 | 119 | 54 | 72 | 14 | 23 | 77 | 13 |
| Male:Female (Male, %) | 284:136 (67.6) | 127:59 (68.3) | 69:50 (58) | 38:16 (70.4) | 47:25 (65.3) | 13:1 (92.1) | 16:17 (48.5) | 52:25 (67.5) | 6:7  (46.2) |
| More than 2 **falls** / year (n, %) | 258 | 97 | 66 | 23 | 40 | 3 | 10 | 41 | 7 |
| Average duration to reach Falls **(>**2 falls/year) milestone Mean ±SD (Months, Range) | 21.1±19.8 (1-140) | 31.2±25.9 (1-120) | 20±16.5 (1-84) | 13.7±13.7 (1-48) | 20.7±18.0 (1-72) | 16.3±17.1 (5-36) | 24.2±15.7 (10-60) | 21.2±17.4 (1-84) | 23.1±16.1 (1-48) |
| Does Subject have significant **cognitive** disability (n) | 71 | 22 | 15 | 7 | 7 | 4 | 3 | 24 | 2 |
| Average duration to reach Significant cognitive disability  Mean ±SD (Months, Range) | 26.3±27.2 (2-150) | 24.2±20.2 (4-60) | 22.4±13.7 (6-48) | 18±20.8 (6-42) | 13±9.9 (6-20) | 15.3±10.2 (4-24) | NA | 24.5±16.7 (8-60) | 15±13 (6-24) |
| Does subject require **urinary** catheter or diapers most of the days (n) | 45 | 17 | 15 | 1 | 3 | 1 | 2 | 6 | 2 |
| Average duration to reach Urinary mile stone (Months, Range) | 27.3±20.1 (4-96) | 50.7±42.4 (1-132) | 36.4±17.5 (6-56) | **NA** | 44.5±30.4 (23-66) | NA | NA | 34.8±20.6 (2-60) | 36±16.9 (24-48) |
| Does subject have severe **Dysphagia** or PEG offered (n) | 25 | 5 | 2 | 3 | 2 | 1 | 4 | 5 | 1 |
| Average duration to reach Dysphagia Milestone (Months, Range) | 32.5±34.1 (6-140) | 24±25 (6-60) | 36±17 (24-48) | **NA** | NA | **NA** | 30.3 ±27 (7-60) | 44.2±32.1 (12-80) | NA |
| Does subject have unintelligible **speech** (n) | 77 | 27 | 17 | 8 | 10 | 6 | 5 | 10 | 1 |
| Average duration to reach Speech milestone (Months, Range) | 27±24.3 (2-120) | 22.6±15.1 (3-48) | 39.9±20.8 (12-84) | 40.7±29.2 (7-60) | 28.5±11.3 (12-36) | 19.8±12.6 (5-36) | 40±18.3 (24-60) | 29.9±19.9 (8-72) | NA |
| Does subject have severe dependency for **mobility** or requires wheel chair (n) | 122 | 36 | 33 | 10 | 14 | 2 | 6 | 16 | 3 |
| Average duration to reach mobility milestone (Months, Range) | 27.7±19.7 (4-120) | 34.8±22.3 (6-90) | 34±17.5 (6-84) | 18.2±18.3 (6-50) | 29.4±20.4 (3-60) | 22±19 (8-36) | 29.4±24.0 (3-60) | 32±21.2 (2-86) | 32±18.3 (12-48) |
| Duration of symptoms when full time care giver was recruited (months)  Mean ±SD  (range) (n) | 26.8±15.9 (4-72) (n=85) | 40.4±28.1 (5-132) (n-23) | 35.6±18.3 (3-84) (n=24) | 36±19.8 (22-50) (n=2) | 36.1±23.3 (3-72) (n=3) | 18±15.9 (6-36) (n=3) | 21.3±23.4 (4-48) (n=3) | 34.9±21.2 (2-84) (n=17) | 42±8.8 (36-48) (n=2) |
| CGI (n-994)   1. Extremely Ill 2. Severely ill 3. Markedly ill 4. Moderately ill 5. Mildly ill 6. Borderline ill 7. Almost normal 8. Not Assessed/ Documented | 1  17  52  124  40  25  38  132 | 0  47  19  41  15  12  3  49 | 2  15  25  28  7  4  2  36 | 0  8  6  15  2  6  7  17 | 0  19  6  19  4  3  0  21 | 0  2  1  4  1  1  0  5 | 0  4  0  8  2  2  0  7 | 1  11  10  22  5  1  0  27 | 0  3  1  7  1  0  0  1 |

*****CGI – Clinical Global Impression of severity *NA : Not available/applicable
