## Appendix for "Progressive Supranuclear Palsy in India: Insights from a Large Multicenter Clinical Cohort (Project PAIR-PSP)"

### Supplementary Appendix – PAIR-PSP Study Protocol

Study Conduct and Methodological Framework of the PAIR-PSP Project

(Pan-India Registry of Progressive Supranuclear Palsy)

**1. Overview and Rationale**

The Pan-India Registry of Progressive Supranuclear Palsy (PAIR-PSP) was created to address the major gap in systematically collected natural history data on PSP within the Indian population. Initially conceptualized as an investigator-initiated project in 2021–2022, PAIR-PSP grew into a coordinated multicenter national study through the Parkinson’s Research Alliance of India (PRAI). From late 2024 onward, the study received formal sponsorship through the Global Parkinson’s Genetics Program (GP2), enabling expansion of recruitment and genotyping. The registry integrates high-quality clinical phenotyping, optional biospecimen and genetic analysis, longitudinal tracking, and a national multi-tier collaboration across clinical sites, a central genetic laboratory, and a dedicated CRO.

**2. Study Governance Structure**

2.1 Steering Committee (PRAI)

A group of senior movement disorder neurologists developed the scientific rationale, protocol, and operational blueprint. Responsibilities included protocol development, selection of participating centers, approving amendments, supervising data quality, and overseeing adjudication.

2.2 CRO Coordination

A third-party Clinical Research Organization (CRO) facilitated operational execution, including site activation, regulatory documentation, IRB tracking, oversight, and communication between clinical centers, MedGenome laboratories, and GP2.

2.3 Genetic Laboratory Partnership

MedGenome India functioned as the central genetic laboratory, overseeing sample receipt, DNA extraction and quality control (QC), genotyping, Bioinformatics and secure data storage on India-based servers only.

2.4 Sponsor

Initially investigator-initiated, the study transitioned to partial sponsorship under GP2 from late 2024, supporting genotyping and expanded infrastructure.

**3. Ethical Framework and Compliance**

3.1 Institutional Ethics Approvals

Each participating center obtained independent IRB/IEC approval before recruitment. Centers joined in phased waves depending on institutional timelines.

3.2 BIORRAP Registration

The project was registered with BIORRAP (ID: PAR23102024), ensuring compliance with Indian biospecimen governance.

3.3 Data and Sample Localization Requirement

All samples, clinical data, and genetic data remained strictly in India. All servers, including Electronic Data capture (EDC) and genetic data servers, were physically located in India.

**4. Participating Centers and Site Selection**

Centers were selected if led by a movement disorders specialist or neurologist with movement disorders interest, and based on feasibility to perform structured data collection, including optional biospecimen modules.

**5. Study Design**

5.1 Type

Prospective, observational, multicenter PSP registry with open-ended recruitment and optional longitudinal follow-up (annual recommended).

5.2 Population

Participants meeting MDS-PSP 2017 diagnostic criteria. Optional inclusion of at-risk family members and Healthy controls.

5.3 Timeline

• Investigator-initiated phase: 2021–2022

• Multicenter expansion: 2022–2023

• Sponsorship and genetic arm activation: From late 2024 onward

**6. Recruitment and Enrollment Workflow**

1. IRB approval at site.

2. CRO site activation and training.

3. Screening with MDS-PSP 2017 criteria.

4. Written informed consent.

5. Baseline assessments.

6. Data entry into HIPAA-compliant India-based cloud EDC.

7. Automated Unique ID generation (PAIR-PSP-XXXX).

8. Biosamples labeled with the Unique ID.

9. Sample shipment to MedGenome.

10. Genetic workflow (DNA extraction, QC, genotyping, Bioinformatics).

**7. Data Capture and Management**

7.1 EDC System

All data were stored in secure India-based servers. Site PIs accessed only their own participants. PRAI Core, CRO, and MedGenome accessed data as per predefined access levels.

7.2 Data Validation

Mandatory fields, logic checks, and periodic audits ensured accuracy and minimized missing fields.

**8. Clinical Battery and Variables**

8.1 Minimum Dataset

Demographics, diagnostic confirmation, phenotype form, PSPRS, treatment history, and essential MDS-PSP 2017 diagnostic features.

8.2 Extended Dataset (Optional)

PSP-CDS, UPDRS III, NMSS, MOCA, FAB, PSP-QOL, Zarit Burden, CGI-S/CGI-C, Treatment received / outcomes, imaging (clinical only), biosamples, and adverse effects.

**9. Biosample Processing and Genetic Workflow**

Blood samples labeled with Unique ID were transported under CRO supervision to MedGenome.

Genetic workflow included:

- DNA QC
- Genotyping via Illumina.
- Gender prediction and relatedness QC
- Genotype rate QC (>99% success)
- Heterozygosity checks
- Ancestry determination (PCA, UMAP, ADMIXTURE)
- Batch effect evaluation
- Outlier identification and dataset finalization
- Additional genetic analysis, structural and functional analysis as required basis.

**10. Quality Control Workflow**

10.1 Clinical QC

Three independent neurologists reviewed all entries, checked for missing or inconsistent data, contacted site PIs for clarification, and held consensus adjudication meetings. Unresolvable data were excluded.

10.2 Genetic QC

Included sex mismatch detection, relatedness, identity confirmation, outlier removal, and creation of a high-confidence final dataset.

10.3 Final Data Lock

Merged clinical + genetic dataset using the Unique ID. No identifiable information was shared between systems.

**11. Longitudinal Follow-up**

Annual assessments included PSP-CDS/PSPRS, motor/non-motor worsening, cognitive decline, ADL burden, caregiver burden, and medication changes. Loss to follow-up was documented with “End of Study” modules.

**12. Data Security and Sovereignty**

12.1 India-Only Data Storage

All EDC data and all genetic data (raw and processed) were stored exclusively on India-based encrypted servers. No samples or data left India.

12.2 Confidentiality

Role-based access, encrypted transfers, pseudonymization via Unique ID, and CRO-governed audits ensured full compliance with regulatory standards.

**13. Replication Framework for Future Studies**

PAIR-PSP establishes a scalable, reproducible model for rare disease registries in India. Strong governance, CRO-guided execution, tiered data capture, structured QC, and India-based data localization make the model adaptable for broader movement disorder research.

**14. Summary**

PAIR-PSP is a landmark multicenter Indian initiative integrating rigorous clinical phenotyping, longitudinal follow-up, standardized biospecimen workflows, high-quality genetic QC, and strict India-only data localization. The collaboration of PRAI, CRO, MedGenome, and GP2 has created one of the most comprehensive PSP datasets from Asia, contributing meaningfully to global understanding of PSP biology and phenotype diversity
